# MRI improves global outcome prediction after moderate or severe traumatic brain injury

**DOI:** 10.64898/2026.10.03.26363950

**Authors:** Hui Shi, William H. Curley, Martina Gjyzari, Ajitesh Nanda, Yelena G. Bodien, Xiaoying Sun, Jacob L. Stubbs, Edilberto Amorim, Nancy R. Temkin, Pratik Mukherjee, Karl A. Zimmerman, Guido Bertolini, Sandra Magnoni, Vincent Dunet, Mauro Oddo, Federico Moro, Emma-Jane Mallas, Neil S. N. Graham, David J. Sharp, Geoffrey T. Manley, Sonia Jain, Michael D. Fox, Brian L. Edlow, Samuel B. Snider, the TRACK-TBI Investigators

## Abstract

**Importance:** Brain MRIs are routinely acquired following moderate or severe traumatic brain injury (TBI) because of their high sensitivity for detecting traumatic brain lesions. However, there is no standard framework for their interpretation, and their clinical utility is unproven.

**Objective:** Here, we test whether MRI-based quantification of traumatic brain lesions improves outcome prediction relative to the best tools currently available to clinicians.

**Design, Setting and Participants:** We analyzed hemorrhagic contusions and axonal injury in four prospective studies of moderate or severe TBI: BIOmarkers of AXonal injury after traumatic brain injury (BIOAX) 2017-2021; Resting and Stimulus-based Paradigms to detect Organized NetworkS and predict Emergence of consciousness (RESPONSE Pilot and RESPONSE 2) 2012-2022; Transforming Research and Clinical Knowledge in Traumatic Brain Injury (TRACK-TBI) 2014-2020. External validation in a fifth, prospective single-center study (Brigham and Women’s Hospital) 2024-2026.

**Exposure:** Contusion volume was measured via manual segmentation. Axonal injury volume was quantified with diffusion tensor imaging, by measuring the volume of skeletonized fractional anisotropy significantly lower than that of healthy controls.

**Main Outcomes and Measures:** Functional outcomes were assessed at 6 months with the Glasgow Outcome Scale-Extended; values 4 were considered unfavorable.

**Results:** Among 201 patients with MRI (mean age: 41, median Glasgow Coma Scale [GCS] score on admission: 7, MRI obtained median 14 days post-injury), MRI-measured contusion volume discriminated outcomes better than CT Marshall score (AUC 0.81 vs 0.64; p=0.001), and remained associated with unfavorable outcomes controlling for multiple commonly-measured clinical variables. Quantitative axonal injury volume discriminated outcomes better than manual MRI grading (AUC 0.82 vs 0.62; p<0.001) and remained associated with unfavorable outcomes independent of other clinical predictors. The prognostic discrimination of traumatic lesion volumes was not improved by accounting for the location or normative connectivity of either lesion type. In a multivariate model, contusion and axonal injury volumes were independently predictive, with age-dependent volume thresholds for 90% probability of unfavorable outcome, and strong discrimination in an external cohort (N=27; AUC 0.83, p<0.001). Replacing all CT-based measures with MRI lesion volumes in the IMPACT model significantly improved out-of-sample prognostic performance (AUC 0.85 vs 0.64; p<0.05).

**Conclusions and Relevance:** MRI-based lesion quantification improves prognostic discrimination relative to measures currently available to clinicians treating patients with TBI. Global functioning at 6 months is driven more by the volume than the location or connectivity of traumatic lesions. These findings support integrating MRI-based lesion quantification into clinical workflows and updated TBI classification frameworks.

## Introduction

Traumatic brain injury (TBI) is among the most prevalent and disabling neurological conditions worldwide. Heterogeneity of the underlying injuries is the central challenge for clinicians providing prognostic assessments to families and for researchers pursuing uniformity in clinical trials. Existing TBI classification frameworks and prognostic models rely exclusively on clinical characteristics at hospital admission and CT-based measures, reflecting the data available at the time these approaches were developed.^1–3^ Yet these measures only indirectly characterize the underlying brain injury. Clinical prognostication therefore remains imprecise, with wide hospital-level variation in rates of withdrawal of life-sustaining therapy,^4,5^ and clinical trials are challenged by heterogeneity, enrolling patients with markedly different injury patterns, burdens, and outcomes. Recognizing these limitations, a recent international initiative has called for an updated TBI classification approach, with imaging-based pathoanatomical features comprising one of four proposed “pillars.”^20^

MRI has the potential to address both challenges by directly characterizing the underlying traumatic brain injury, thereby improving clinical prognostication and enabling biologically informed patient stratification for clinical trials. Compared with CT, MRI is more sensitive for detecting the two major forms of structural injury after TBI: hemorrhagic contusions and axonal injury.^6–8^ Hemorrhagic contusions, heterogeneous lesions caused by contact between brain parenchyma and the skull surface, can be visualized using conventional structural MRI sequences.^6,9,10^ Axonal injury, the swelling and and/or disruption of white matter fibers,^11^ can be identified using T2-weighted, susceptibility-weighted, and diffusion tensor imaging (DTI) sequences.^12–15^ Together, these measurements enable characterization of the extent and distribution of traumatic brain injury in vivo.

Despite its promise, MRI is not yet a standardized tool for TBI prognostication or classification. Although MRI is increasingly acquired clinically after moderate or severe TBI for qualitative assessment of injury,^16,17^ identification of traumatic lesions – particularly axonal injury – can vary across readers.^19^ Quantitative approaches, including DTI, may improve the reproducibility of injury measurement but remain largely confined research settings.^18^ More fundamentally, there is no established framework for translating MRI-defined traumatic lesions into estimates of functional recovery.^4,5^ An important unresolved question is whether outcomes are better explained by the location or volume of traumatic brain lesions. Some studies have linked outcomes to injuries involving specific brain regions,^21–25^ whereas others suggest that global lesion volume is the stronger predictor.^26–28^ Clarifying the relative importance of lesion volume and location is essential for developing a standardized, quantitative framework for MRI-based prognostication and patient stratification after TBI.

Here, we analyzed data from four prospective studies of moderate or severe TBI conducted in Europe and the United States to test specific hypotheses about the prognostic value of MRI-measured traumatic lesions. For each lesion type – hemorrhagic contusions and axonal injury – we tested whether MRI-measured lesion volumes were associated with 6-month outcomes independent of other clinical and radiological information, and whether accounting for lesion location improved prognostic discrimination relative to volume alone. We validated our primary findings in a fifth, prospectively collected dataset.

## Materials and methods

### Cohort Characteristics

We formed a Derivation Cohort (N=201 patients, 315 controls) with data from four prospective TBI studies. The original studies included: BIOmarkers of AXonal injury after traumatic brain injury (BIOAX; NCT03534154),^29,30^ a multi-center, European study with data collected between November 2017 to September 2021, Resting and Stimulus-based Paradigms to detect Organized NetworkS and predict Emergence of consciousness (RESPONSE Pilot and RESPONSE 2; NCT03504709),^31,32^ two single-center, American studies, enrolling between June 2012 and December 2022, and Transforming Research and Clinical Knowledge in Traumatic Brain Injury (TRACK-TBI; NCT01565551),^33–35^ a multi-center American study with data collection between March 2014 and August 2020. Detailed descriptions of each study have been previously published. The External Validation Cohort (N=27 patients, 2 controls) was a prospective, single-center study of moderate or severe TBI at the Brigham and Women’s Hospital ([BWH]; Level 1 Trauma Center, Boston MA) enrolling since 2024 (Supplementary Methods).

We included all patients with requisite data in each analysis, all of whom had moderate or severe TBI (at least one Glasgow Coma Scale [GCS]^36^ score < 13 within 24 hours of injury), MRI acquired in the acute or subacute phase, and a 6-month outcome obtained using the Glasgow Outcome Scale-Extended (GOSE),^37,38^ a widely used instrument for assessing global levels of dependency after TBI (Supplementary Methods). Age- and sex-matched controls from each study with no history of TBI were also included for normalization of imaging features. Consort diagrams for each analytic cohort are provided in Supplementary Figure 1.

International Mission for Prognosis and Analysis of Clinical Trials in TBI (IMPACT)^1^ score variables were abstracted from the medical record as part of standard data collection procedures in each study. This included the CT Marshall Score,^39^ a standard, ordinal measure of traumatic injury severity on CT. This was scored on the first head CT after admission (TRACK-TBI) or the latest CT on the first day post injury (BIOAX, RESPONSE, BWH). Command following at time of MRI was determined from behavioral measures – Coma Recovery Scale-Revised for participants from RESPONSE Pilot and RESPONSE 2, and GCS for participants from TRACK-TBI and BIOAX. As only total GCS scores were recorded in BIOAX, patients with total scores < 8 were considered not to be following commands.

### Image Processing

#### Conventional MRI

Imaging acquisition parameters from each study have been previously published. Areas of hemorrhagic contusion were manually traced in standard fashion on available T1 multi-echo magnetization-prepared rapid acquisition gradient sequence (T1-MPRAGE) and Fluid-Attenuated Inversion Recovery (FLAIR) sequences (Supplementary Methods; Supplementary Figures 2-3). Conventional sequences were manually evaluated for axonal injury by consensus judgement of two raters (S.B.S and W.H.C). Raters screened for characteristic lesions at the grey-white junction, corpus callosum, and brainstem on FLAIR and susceptibility-weighted or gradient echo/T2* sequences that were distinct from hemorrhagic contusions. For participants without either heme-sensitive or FLAIR sequences, only the available sequences were used. Each scan was then scored according to the Adams/Gentry ordinal classification scheme (grade I = lobar white matter involvement, grade II = callosal involvement, grade III = brainstem involvement).^11,44,45^

#### Diffusion Tensor Imaging (DTI)

DTI acquisition parameters from each study are provided in Supplementary Table 1 (A-G) and pre-processing is described in the Supplementary Methods. Adapting approaches previously used in other contexts,^13,51,52^ each participant’s map of white matter integrity (fractional anisotropy [FA]) was skeletonized, with each voxel normalized by the mean and standard deviation among controls (N=315) scanned from the same study on the same scanner (Supplementary Figure 4). For each participant, we defined lesioned voxels along this skeleton as those with FA falling at least two standard deviations below the control mean. This value was selected based on precedent set by prior studies,^53,54^ with the intention of balancing sensitivity with specificity for lesion detection.

### Primary Outcome

The primary outcome was the GOSE at 6 months post-injury (Supplementary Methods). Total scores range from 1 (death) to 8 (asymptomatic).^37,38^ This outcome was treated as a continuous variable for generating brain-wide statistical association maps and was binarized at > 4 (favorable) versus ≤ 4 (unfavorable) for comparative analyses. Participants with GOSE ≤ 4 cannot function independently outside the home. This threshold has been used in multiple randomized controlled treatment trials of TBI.^1,55–58^

### Statistical Analysis

#### Contusion volume, location, and functional outcomes

The area under the receiver operating characteristic curve (AUC) of total MRI-measured contusion volume (cc) for discriminating 6-month favorable from unfavorable outcomes was compared to that of the CT Marshall Score^39^ using Delong’s test. We then used multivariate logistic regression to determine whether MRI-measured contusion volume was independently associated with unfavorable 6-month outcomes, controlling for controlling for GCS_motor_ score on hospital admission, command-following at time of MRI, or both variables. Contusion volume was log-transformed for this analysis given its right-skewed distribution. We next designed a custom pipeline to test whether contusion location or structural/functional connectivity improved discrimination of 6-month unfavorable outcomes relative to contusion volume alone (Supplementary Methods; Supplementary Figure 3).

#### Axonal injury volume, location, and functional outcomes

We compared the AUC of the total volume (ccs) of lesioned white-matter skeleton voxels (FA > 2 SD below control mean) for discriminating 6-month favorable from unfavorable outcome to the AUC of the manually-graded Adams/Gentry classification using Delong’s test. In an exploratory analysis, we also compared the discrimination of region-of-interest-based approaches (as opposed to the TBSS-skeleton) for measuring FA (Supplementary Methods). We used multivariate logistic regression to determine if white matter skeleton lesion volume was independently associated with unfavorable 6-month outcomes, controlling for GCS_motor_ score on hospital admission, command-following at time of MRI, or both variables. White matter lesion volume was log transformed for this analysis given its right-skewed distribution. Lastly, we used a customized pipeline to test whether axonal injury location or connectivity improved the accuracy of outcome predictions relative to axonal injury volume alone (Supplementary Methods; Supplementary Figure 4).

#### Independent contribution of contusions and axonal injury

Among participants with both conventional MRI data and DTI, we used multivariate logistic regression to determine whether (log-transformed) axonal injury lesion volume and contusion volume were independently associated with the likelihood of 6-month unfavorable outcomes. Age (per decade) was included as a biological variable. We predicted probabilities of unfavorable outcomes in the External Validation cohort using the locked multivariate logistic regression model from the Derivation Cohort and defined discrimination with AUC.

#### Integrating MRI-measured lesion volumes into the IMPACT score

To test the hypothesis that MRI-measured lesion volumes improve the best available comprehensive prognostic model, we fit two logistic regression models in the Derivation Cohort. The first model included all variables in the IMPACT (Core+Extended+Lab) model. The second model replaced all CT-measured variables (Marshall Score, traumatic subarachnoid hemorrhage, and epidural hematoma) with log-transformed MRI-measured contusion and axonal injury volumes. Finally, we predicted probabilities of unfavorable outcomes in the External Validation cohort using each of the two models and compared the AUC curves with Delong’s test.

All statistical analyses were performed in R (RStudio Version 2024.12.0+467).

## Results

### Patient Characteristics

The mean (SD) age of the N=201 Derivation Cohort patients who underwent conventional MRI, DTI, or both, was 41 (18). 73% were male and 88% were white (Table 1). The median (IQR) GCS on hospital admission was 7 (3, 10), the most common CT Marshall score was II, 75 (37%) had an unfavorable outcome (GOSE 4) at 6 months post-injury, and 10 (5%) died (GOSE 1). MRI was acquired at a median (IQR) 14 (13, 15) days post injury and 59% of those with available data were following verbal commands at time of MRI acquisition. Demographics and injury characteristics differed by study of origin (Supplementary Table 2). Compared to the Derivation Cohort, External Validation Cohort patients were older, less likely to identify as white, and more likely to have contusions (Supplementary Table 3). They had larger axonal injury volumes, shorter times to MRI, and greater (IMPACT) predicted probabilities of unfavorable 6-month outcome (Supplementary Table 3).

**Table 1.** Derivation Cohort Characteristics s (N=201)

| **Table 1:** Derivation Cohort Characteristics (N=201) |  |
| --- | --- |
| Mean Age (SD) | 41 (18) |
| Male Sex (%) | 146/201 (73%) |
| Race (%) |  |
| White | 171/195 (88%) |
| Black | 15/195 (8%) |
| Other | 9/195 (5%) |
| Study of Origin (%) |  |
| RESPONSE Pilot | 18/201 (9%) |
| RESPONSE 2 | 31/201 (15%) |
| BIOAX | 65/201 (32%) |
| TRACK-TBI | 87/201 (43%) |
| Median Admission GCS (IQR) | 7 (7) |
| Pupils (%) |  |
| Both React | 152/184 (83%) |
| One Reacts | 13/184 (7%) |
| Neither Reacts | 19/184 (10%) |
| CT Marshall Score (%) |  |
| I | 37/186 (20%) |
| II | 84/186 (45%) |
| III | 22/186 (12%) |
| IV | 5/186 (3%) |
| V | 12/186 (6%) |
| VI | 26/186 (14%) |
| Hemorrhagic Contusion (%) | 139/200 (70%) |
| Median Hemorrhagic Contusion Volume (cc, IQR) | 11 (59) |
| Axonal Injury Grade (%) |  |
| 0 | 14 (9%) |
| I | 43 (27%) |
| II | 48 (30%) |
| III | 55 (34%) |
| Mean Scaled White Matter Mean FA (SD) | -1.6 (1.5) |
| Median White Matter Lesion Volume (cc, IQR) | 6.6 (12.8) |
| Median Days to MRI (IQR) | 14 (2) |
| Following Commands at MRI | 71/120 (59%) |
| Mean (mm) Motion During DTI (SD) | 0.93 (0.90) |
| Median IMPACT<sub>Core+CT+Lab</sub> Probability of Unfavorable Outcome (N=146) | 0.37 (0.38) |
| 6-month GOSE ≤ 4 (%) | 75/201 (37%) |

### Cerebral Contusions

Among 200 Derivation Cohort participants with conventional MRI sequences (Supplementary Figure 1), 139 (70%) had hemorrhagic contusions (median [IQR] volume: 11 cc [0, 59]). MRI-measured contusion volume discriminated 6-month outcomes significantly better than a standard, CT-based ordinal scale of injury severity (MRI volume AUC: 0.81 vs Marshall Score: 0.64; p = 0.001; Figure 1A). The incidence of unfavorable functional outcomes increased with greater contusion volume (Figure 1B). MRI-measured contusion volume remained associated with likelihood of unfavorable outcome accounting for admission GCS_motor_ score (aOR 1.6 [1.4, 1.9] per doubling of contusion volume; p < 0.001; Figure 1C), command following at time of MRI (aOR 1.4 [1.2, 1.7]; p < 0.001; Figure 1D), or both variables (aOR 1.4 [1.2, 1.7]; p < 0.001).

**Figure 1:**
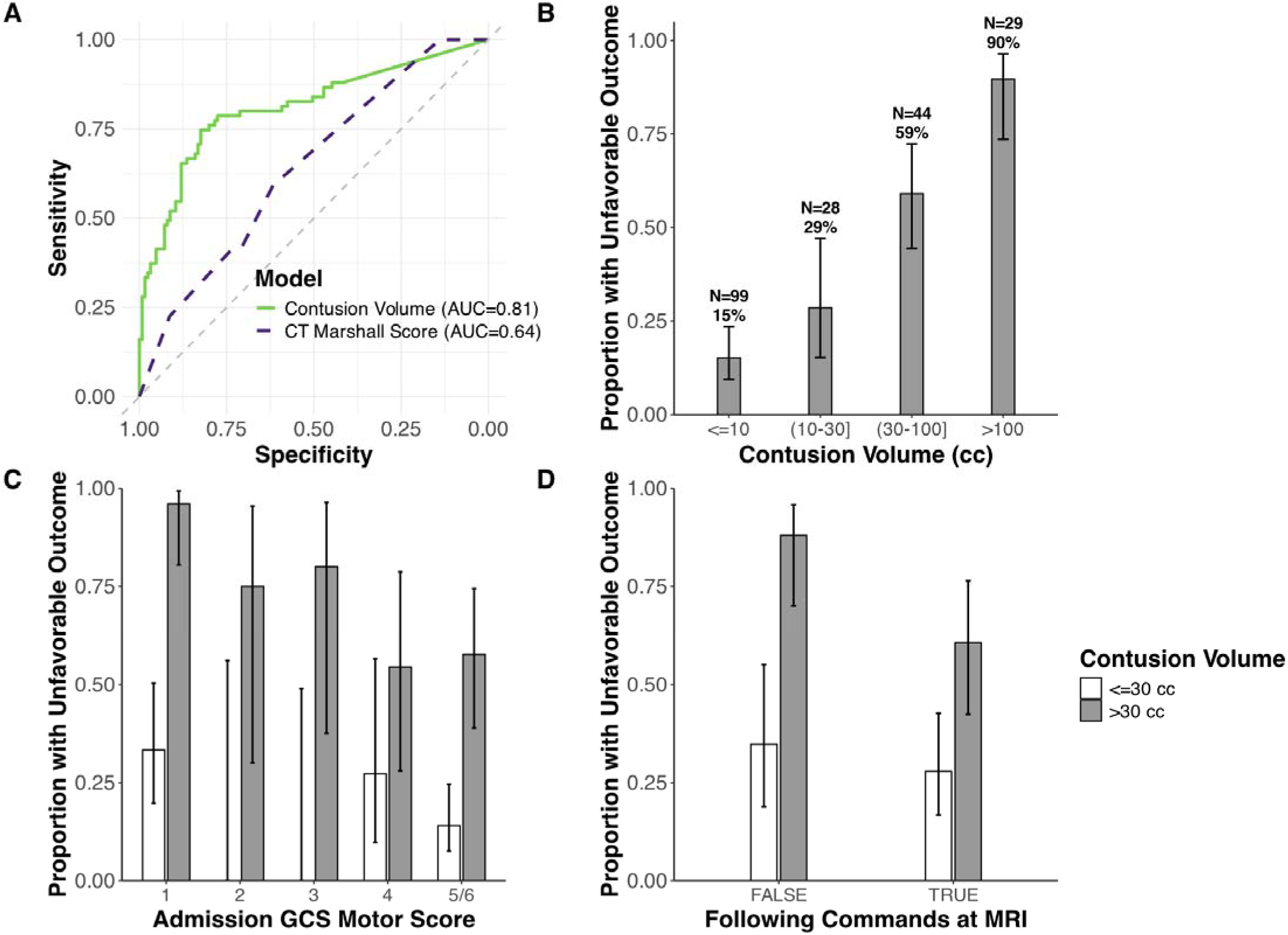
MRI-measured total contusion volume discriminates functional outcomes better than CT Marshall Score. (A) ROC curves of MRI-measured contusion volume (green) and CT Marshall score (purple). Bar graphs of proportion of unfavorable outcome with 95% confidence intervals across contusion volume groupings (B), or stratified by contusions less than or greater than 30 ccs across admission GCS motor scores (C), and command following at time of MRI (D).

Contusions were primarily distributed throughout the inferior frontal and temporal regions (Figure 2A). The strongest regional associations with unfavorable outcome were in the right temporal and left frontal regions (P_FWE_ < 0.05; Figure 2B). Weighting contusion volumes based on degree of overlap with these regions (Damage Score; Supplementary Methods) performed worse at discriminating 6-month outcomes than raw (location-agnostic) contusion volume (Figure 2C; Damage Score AUC: 0.70 vs. volume AUC: 0.81; p = 0.004).

**Figure 2:**
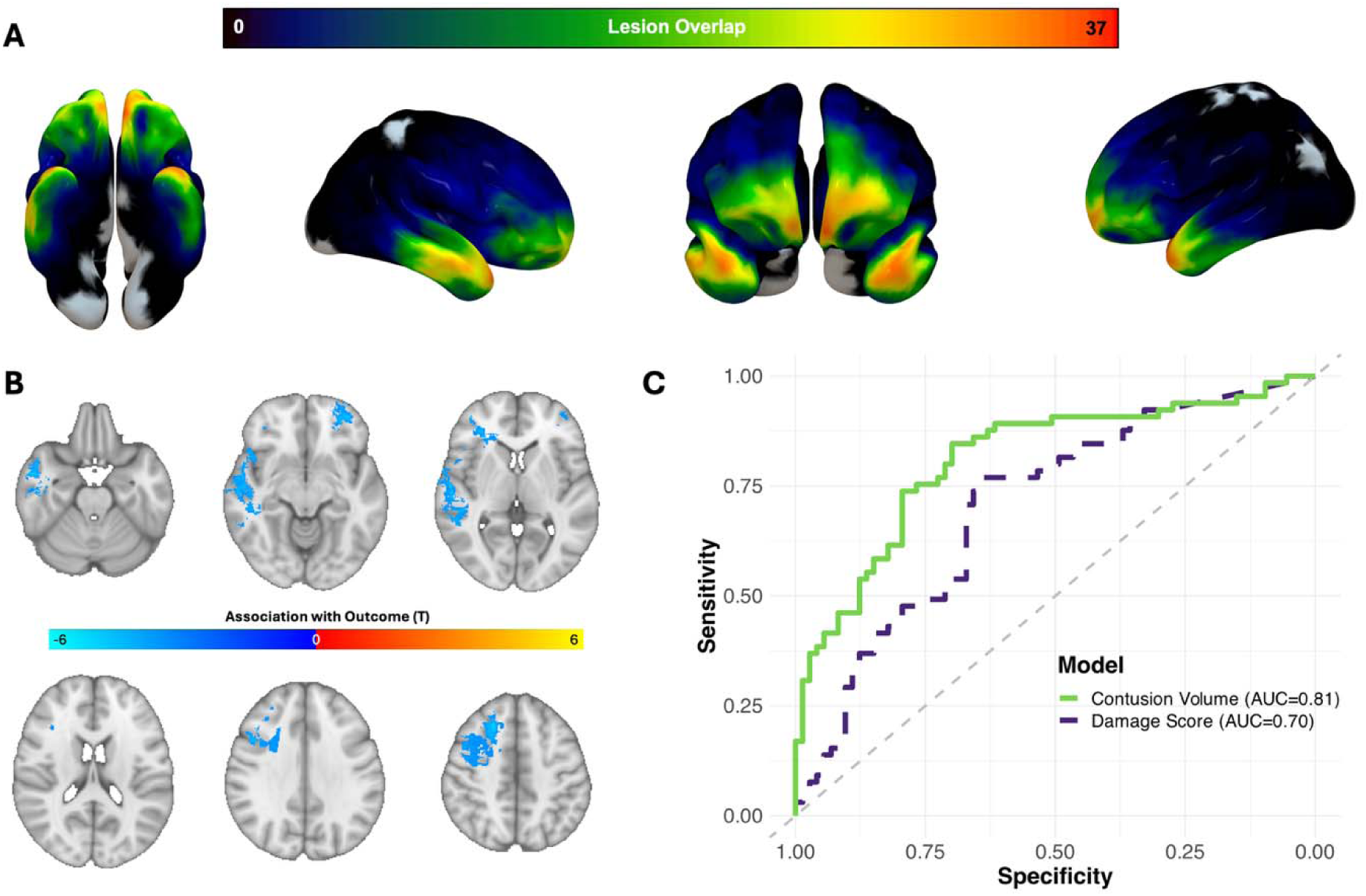
Contusion location does not improve outcome discrimination. (A) Contusion overlap surface map. The color bar indicates the number of participants with lesions at each location. (B) Voxel-outcome association map with 1,000 permutation-based cluster multiple comparisons correction (raw p threshold of 0.001; P_FWE_ < 0.05 voxels are shown). The color bar indicates T values of significant voxels. Areas with negative (blue) T values are those that, when lesioned, result in a lower 6-month GOSE score. (C) ROC curves comparing Damage Score (accounting for lesion location, purple) to location-agnostic total contusion volume (green).

Connectome-informed Damage Scores, which incorporate the normative structural and functional connectivity of each contusion location (Supplementary Methods), did not improve outcome discrimination relative to raw contusion volume (Supplementary Figure 5A-D), nor were they independently associated with outcome after controlling for contusion volume (structural: aOR 1.6 [0.6, 4.9] per 1 SD score increase; p = 0.43]; functional: aOR 1.32 [ 0.7, 2.6 per 1 SD score increase; p = 0.41).

### Axonal Injury

Among 164 Derivation Cohort patients with DTI, 27% had visually-assessed grade I axonal injury, 30% had grade II, and 34% had grade III. Quantified along a standard skeleton and compared against 315 matched controls, the median axonal injury lesion volume was 6.6 cc [3.5, 16.3]. DTI-measured axonal injury lesion volume discriminated 6-month outcomes significantly better than visual inspection-based grading (lesion volume AUC: 0.82 vs manual: 0.62; p < 0.001; Figure 3A), as well as region-of-interest based measures of mean white matter FA (Supplementary Methods; Supplementary Figure 6). The incidence of unfavorable 6-month functional outcomes increased with greater axonal injury volume (Figure 3B). MRI-measured axonal injury volume remained associated with likelihood of unfavorable outcomes accounting for admission GCS_motor_ score (aOR 3.3 [2.2, 5.0] per doubling of axonal injury volume; p <0.001; Figure 3C), command following at time of MRI (aOR 2.7 [1.7, 4.6]; p < 0.001; Figure 3D), or both variables simultaneously (aOR 2.8 [1.6, 4.7]; p < 0.001).

**Figure 3:**
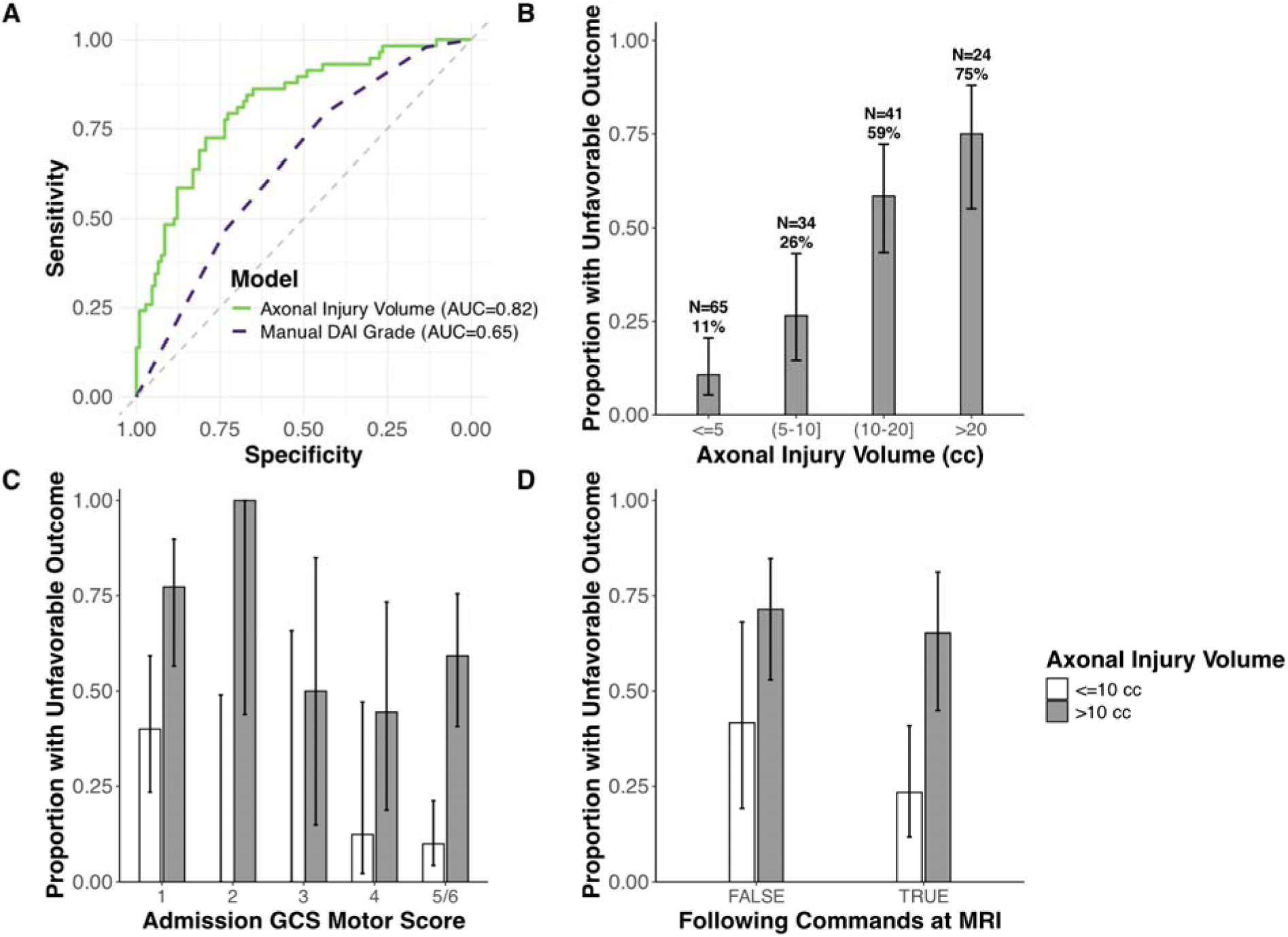
MRI-measured axonal injury volume discriminates functional outcomes better than visual inspection-based Adams/Gentry grading scale. (A) ROC curves of white matter skeleton lesion volume (green) and visual inspection-graded diffuse axonal injury (DAI) severity (Adams/Gentry grading, purple). Bar graph with 95% confidence intervals comparing the proportion of unfavorable outcome in each group of skeleton lesion volume (B), stratified by axonal injury volume less than or greater than 10 ccs across admission GCS motor scores (C), and command following at time of MRI (D).

Most axonal injury occurred in the corpus callosum, followed by the temporal white matter and cingulum (Figure 4A). The strongest regional associations with outcome were observed in the same regions as well as the internal and external capsules and the cerebral peduncles (P_FWE_ < 0.05; Figure 4B). Weighting axonal injury lesion volumes based on degree of overlap with these regions (Damage Score; Supplementary Methods) did not improve outcome discrimination relative to axonal injury lesion volume alone (Figure 4C; Damage Score AUC= 0.78 vs. volume AUC 0.82; p = 0.09). Structural connectivity-informed Damage Scores did not improve outcome discrimination relative to axonal injury lesion volume alone (Supplementary Figure 7).

**Figure 4:**
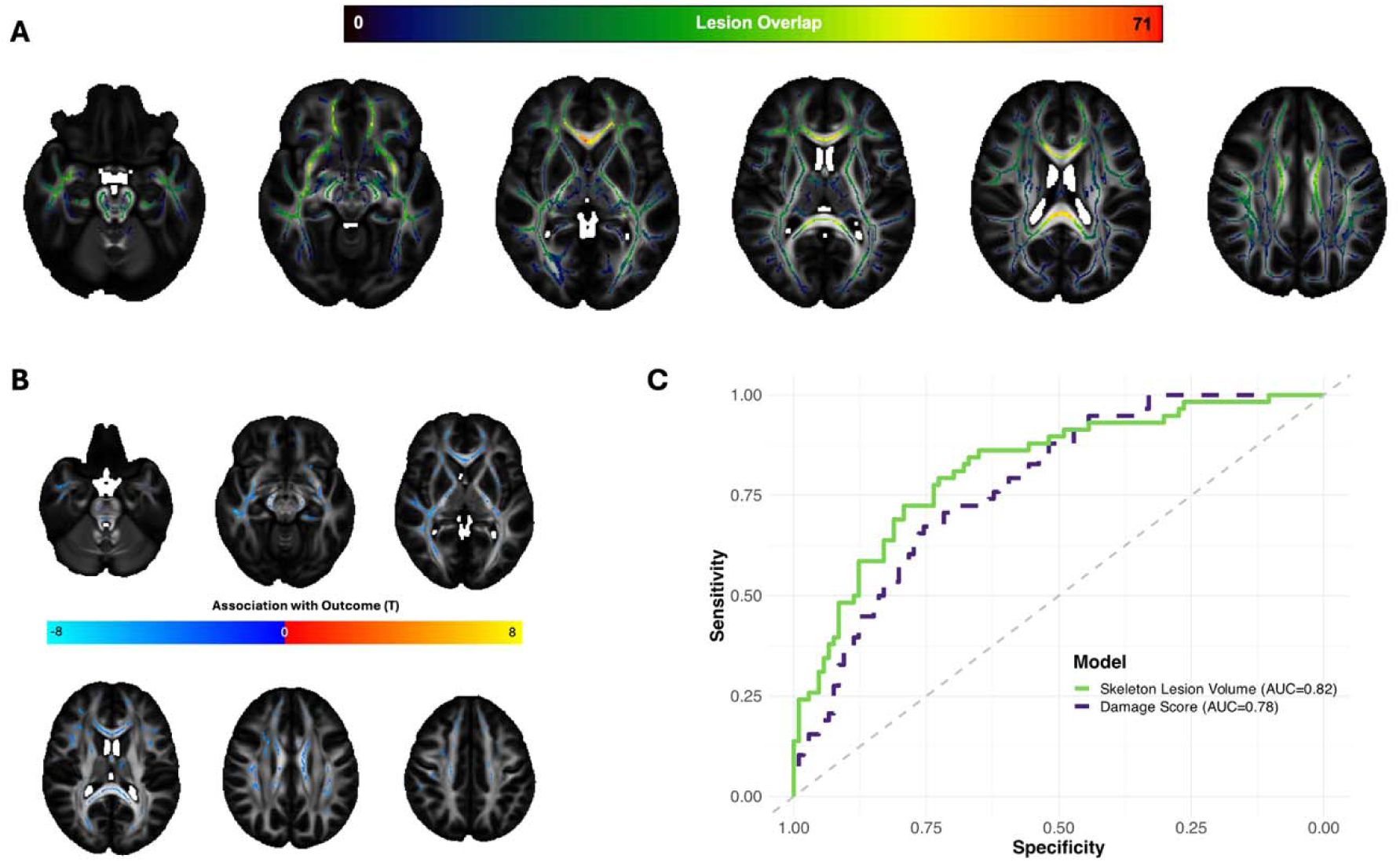
Axonal injury location does not improve outcome discrimination. (A) Lesions along a white matter skeleton are overlaid on a standard FA map in MNI space. The color bar indicates the number of participants with lesions at each point on the skeleton. (B) Voxel-outcome association map with permutation-based cluster multiple comparisons correction (raw p threshold of 0.001; P_FWE_ < 0.05 voxels are shown. The color bar indicates T values of significant voxels. Areas with negative (blue) T values are those that, when lesioned, result in a lower 6-month GOSE score. (C) ROC curves comparing the discriminative ability of location-based Damage Score (purple) to that of global axonal injury lesion volume (green).

### Prognostic Independence and External Validity of MRI-Measured Lesion Volumes

In a combined model together with age, contusion volume and axonal injury volume were independently associated with unfavorable outcomes (Supplementary Table 4). To demonstrate the joint contribution of contusion and axonal injury lesion volumes to likelihood of 6-month unfavorable outcomes, we map model predictions for patients of four different ages in Figure 5A, with calibration shown in Figure 5B. Exceeding a 90% predicted probability of unfavorable outcome (Figure 5A; beyond dashed white line) required enormous traumatic lesion volumes in 25-year-old patients (∼ 300 ccs contusion volume and ∼ 60 ccs axonal injury volume), and more modest lesion volumes (∼30 ccs contusion volume and ∼ 20 ccs axonal injury volume) in 75-year-old patients.

**Figure 5:**
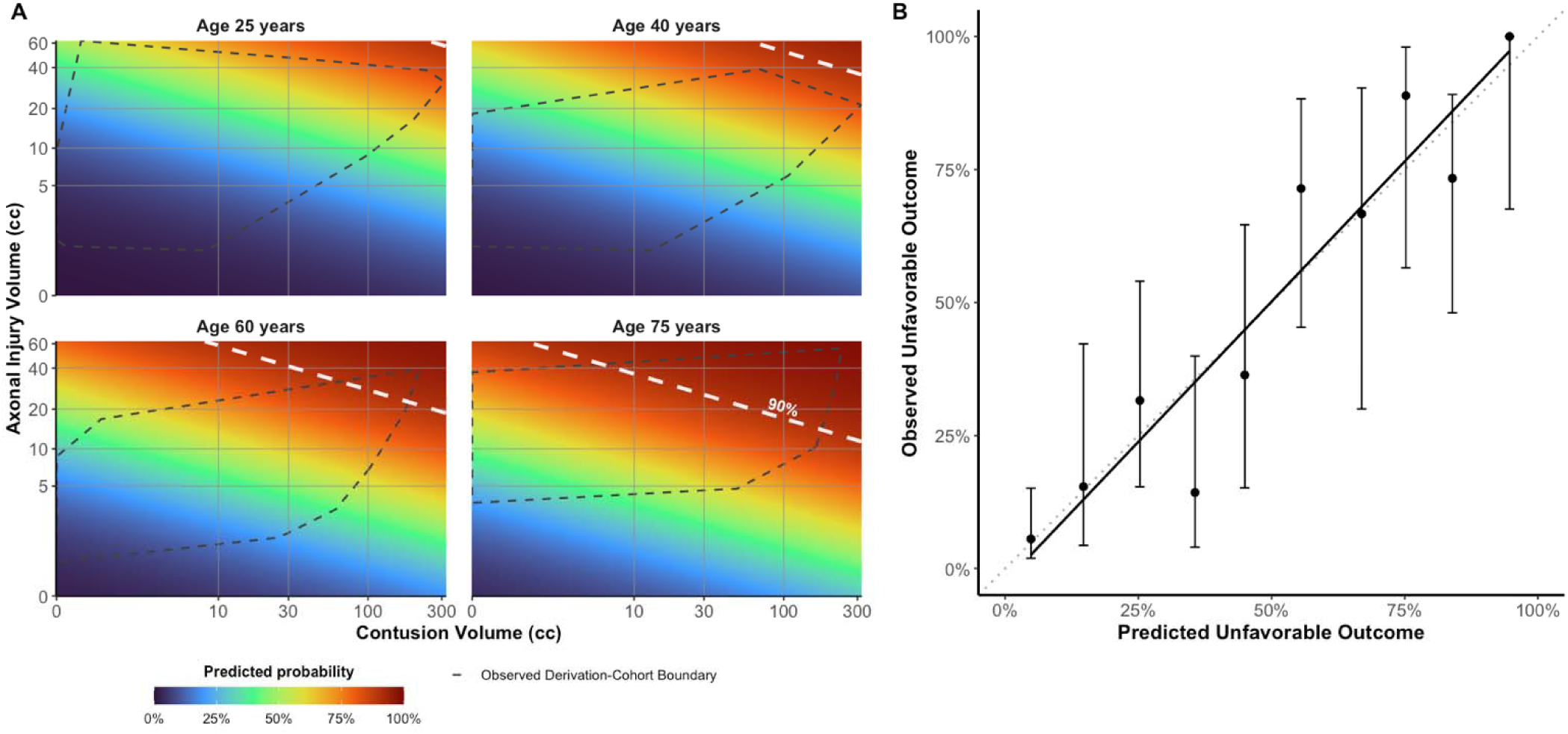
MRI-measured contusion and axonal injury volume independently predict outcome with age-dependent thresholds. (A) Heatmap of the multivariable model predicted probability of unfavorable outcome by volume of contusion and axonal injury, stratified by age. Dashed white line represents threshold for > 90% probability unfavorable outcome. Dashed black line represents the boundary of observed patient characteristics within the Derivation Cohort. (B) Calibration plot of ten predicted probability groupings (x axis) and true observed proportion of unfavorable outcomes (y axis) with 95% confidence interval.

As observed in the Derivation Cohort, there was a progressive increase in the incidence of unfavorable 6-month outcomes with increasing lesion volumes in the External Validation cohort (Figure 6A, B). Multivariate model-predicted probabilities of unfavorable 6-month outcome demonstrated strong discrimination between outcome groups (AUC 0.83 [0.67, 0.98]; p < 0.001; Figure 6C, D).

**Figure 6:**
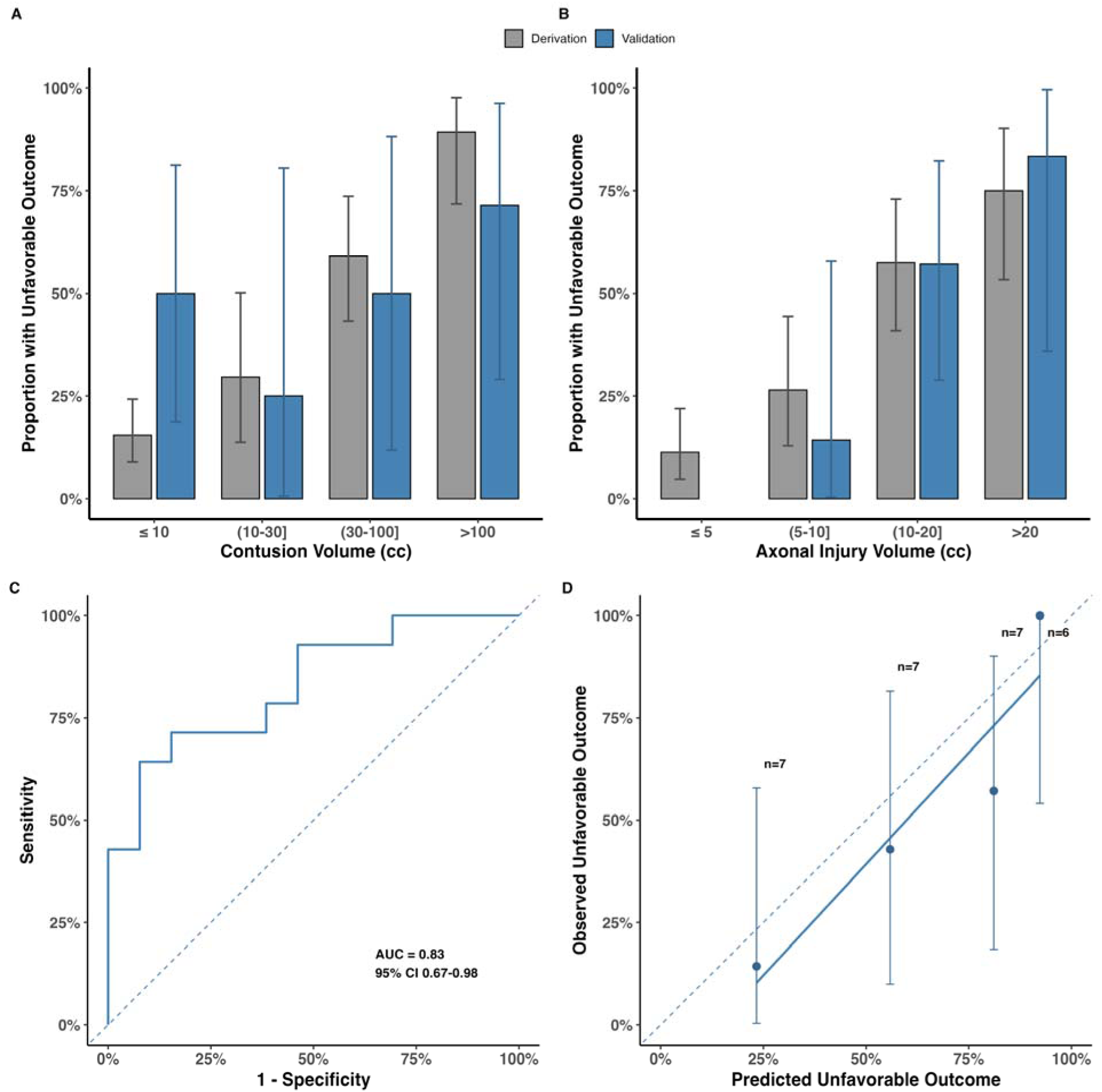
Contusion and axonal injury volumes accurately discriminate outcomes in external validation cohort. Bar graphs with 95% confidence intervals comparing the proportion of unfavorable outcome in each bucket of contusion volume (A) and axonal injury volume (B) for both derivation (gray) and external validation (blue) cohorts. (C) ROC curve for multivariate model-prediction of poor outcome in external validation cohort. (D) Calibration plot of four predicted probability groupings (x axis) and true observed proportion of unfavorable outcomes (y axis) with 95% confidence intervals.

Finally, we compared the performance of the full IMPACT (Core+Extended+Lab) model to a new model replacing CT-variables with MRI lesion volumes (IMPACT_Core+MRI+Lab_). In the External Validation Cohort, the discriminative performance of an IMPACT model including MRI lesion volumes was superior to the full IMPACT model (AUC 0.85 vs 0.64; p = 0.046; Supplementary Figure 8).

## Discussion

In one of the largest pooled analyses of MRI in moderate and severe TBI, including more than 200 patients across two continents and more than 10 medical centers, MRI-quantified volumes of contusion and axonal injury provided prognostic information beyond established clinical and radiological measures. Both lesion types independently contributed to the likelihood of unfavorable 6-month functional outcomes, with tolerance thresholds that varied by age and replicated in an independent, prospectively-collected cohort. In contrast, incorporating lesion location or brain-wide connectivity did not improve prognostic discrimination beyond volume alone. Collectively, these findings suggest that quantitative MRI measurement of traumatic lesion burden may provide a simple and generalizable means of incorporating pathoanatomical injury into TBI prognostication and classification. MRI-measured traumatic lesion volume warrants consideration as a candidate for the imaging pillar of the recently proposed multidimensional TBI classification framework.^20^

There is currently no established framework for the prognostic interpretation of hemorrhagic contusions. While some studies have concluded that the volume of hemorrhagic contusions is associated with likelihood of unfavorable outcome,^28,63^ others have suggested that volume alone is not a robust^64^ or independent^65,66^ predictor. Early studies led to the inclusion of intracranial hemorrhages larger than 25 ccs as an element of the Marshall Score,^39^ a CT classification scheme used in the most widely-cited TBI prognostic model (IMPACT).^67^ However, the discriminative accuracy of the Marshall Score is now contested,^65,68,69^ and the Marshall score was dropped from a second, widely-cited prognostic model (CRASH).^2^ Our work demonstrates the superior discriminative accuracy of MRI-measured total contusion volume relative to the CT-based Marshall score. While we did not directly compare CT-measured contusion volume to MRI-measured contusion volume, the robustness of the association across multiple studies and controlling for multiple covariates suggests that MRI-based measurements may better delineate the full extent of parenchymal injury.

Prognosticating in individual patients based on contusion volume alone requires caution. The likelihood of unfavorable outcome changes only marginally between 0 and 30 ccs of contusion volume, the size of roughly half the contusions in both cohorts. In the absence of significant axonal injury volumes, exceeding 90% probability of unfavorable outcome requires more than 100 ccs of contusion in a 75-year-old patient, and even larger contusions in younger patients. As such, MRI-quantified contusion volume is informative for prognosis, but should be interpreted in the context of age, axonal injury burden, and other patient-specific factors.

In current clinical practice, axonal injury severity is graded (Adams/Gentry) from visual inspection of a diffusion-weighted, T2-weighted, and/or heme-sensitive MRI sequence.^11,44^ Higher grades are associated with higher likelihood of unfavorable outcomes.^70–72^ However, severity grades based on visual inspection have been shown to have limited inter-rater reliability across radiologists.^19^ Furthermore, microhemorrhages on MRI, commonly interpreted as markers of axonal injury, are unreliable indicators of histopathologically-defined axonal injury at autopsy.^73–75^ DTI enables direct quantification of white matter integrity. Several studies have identified associations between DTI-derived evidence of axonal injury and outcomes after moderate or severe TBI.^26,76–80^ Despite prior positive studies, there are two main reasons why DTI is not routinely acquired within standard clinical radiology workflows in patients with TBI. First, prior studies have used a wide variety of approaches to processing and analyzing the data without demonstrated superiority of one approach over another. Second, it has not been clearly demonstrated that DTI adds prognostic information independent from conventional MRI sequences.

Building on previously described techniques,^13^ our fully automated approach to quantifying axonal injury volumes demonstrated superior prognostic discrimination relative to Adams/Gentry grading on conventional sequences (Figure 3A), other automated DTI-based approaches (Supplementary Figure 6), and location/connectivity-informed approaches (Figure 4, Supplementary Figure 7). It yielded groups of axonal injury severity that were prognostically independent from other radiological and clinical information (Figure 5B-D). Most importantly, these predictions generalized to an independent dataset with multiple differences from the Derivation Cohort, including a distinct DTI acquisition sequence and a different scanner (Figure 6). These results highlight a path towards integration into standardized clinical workflows.^13^

As with contusions, we identified an increasing likelihood of unfavorable 6-month outcomes with increasing volume of axonal injury (Figure 3B). Despite this trend, only 75% of patients with the most severe axonal injury volume (> 20 ccs of lesioned white matter skeleton) had unfavorable outcomes – a probability insufficient to convey a uniformly grim prognostic assessment. In a 25-year-old patient, even 60 ccs of axonal injury volume did not produce > 90% probability of unfavorable 6-month outcome. Together with the findings for contusion volume, these results highlight the need to contextualize the volume of each lesion type with other injury-related and patient-specific factors.

Predictions based on patient age and MRI-measured traumatic lesion volumes robustly discriminated favorable from unfavorable outcomes and appeared well-calibrated in external data (Figure 6). Furthermore, the use of MRI rather than CT-based imaging features in the full IMPACT prognostic model demonstrated superior prognostic discrimination in external data. These findings strongly support the integration of MRI-measured traumatic lesion volumes into a multi-dimensional classification framework that includes clinical characteristics, biomarkers, and modifying factors.^20^ Stratifying observational or interventional studies using this new framework will enable enriched clinical trials with more homogeneous injuries and outcome distributions, particularly those targeting the post-acute recovery phase.

Given well-established links between focal brain lesions and specific neurological deficits,^62,83–85^ the lack of prognostic benefit from measuring the location of hemorrhagic contusions and axonal injury was surprising. These findings should not be interpreted to suggest that lesion location is biologically unimportant. Rather, for predicting a multidomain global outcome scale (GOSE), total traumatic lesion volume may carry more prognostic information that the precise spatial distribution of those lesions. Additionally, traumatic brain lesions are distributed in a spatially restricted manner. Contusions tend to occur in inferior cerebral regions where the rough floor of the skull disrupts the surface of the brain parenchyma. The rotational forces that cause traumatic axonal injury preferentially injure white matter regions rendered susceptible to this force based on proximity to structures of differing densities.^89–91^ Within this limited subset of brain regions, total lesion volume may better reflect the likelihood of recovery than injury to any specific structure. Focal lesions in regions previously reported to have independent prognostic significance (e.g., brainstem, substantia nigra, mesencephalon)^22–25,92^ may serve as a proxy for a greater overall burden of traumatic injury.

## Limitations

Important limitations must be considered when interpreting these results. First, additional recovery can occur beyond 6 months in patients recovering from severe TBI.^96,97^ Second, although only ten patients in the Derivation Cohort died before the 6-month assessment point, including any patients in the unfavorable outcome group who died can increase the risk of self-fulfilling prophecy bias. Third, our power to identify specific neuroanatomical associations with outcome was reduced by the limited area of lesion overlap as well as anatomical distortions that may reduce registration accuracy. Fourth, MRIs in the Derivation Cohort were acquired a median of 14 days post-injury, a timepoint too late to inform acute medical decisions. However, most patients with moderate or severe TBI are able to undergo MRI before this time point, and even at two weeks post-injury, many consequential decisions remain. Fifth, our broad definition of moderate or severe TBI (any GCS < 13 within the first 24 hours of injury) may have led to the inclusion of some participants who were classified as mild by standard criteria. Lastly, several additional factors may prevent rapid translation of these findings. Manual contusion tracing is labor intensive and subjective. It is unclear whether the entirety of the mixed density contusion, or just some fraction of it, will evolve into an encephalomalacic cavity. It is additionally unclear how to classify other types of pathology that are found in TBI and may be related to outcome (e.g., diffusion restriction resulting from brain compression). It remains to be seen whether manual tracing is superior to cruder but faster approaches (e.g., the ABC/2 formula).^82^ While the DTI-based analysis is fully automated, it requires setting up a customized pipeline, which may be challenging to integrate into radiological workflows.

## Conclusions

MRI-based lesion quantification outperforms standard imaging-based prognostic frameworks in moderate and severe TBI, has independent prognostic significance relative to other clinical variables, and accurately predicts outcomes in independent data. Global levels of function 6 months after moderate or severe TBI are primarily driven by the volume rather than the location or connectivity of traumatic lesions. This study presents a roadmap for translating MRI findings into prognoses after TBI and identifies a promising candidate for the “imaging pillar” of the newly proposed TBI reclassification framework.^20^

## Data availability

The data used in this study originates from multiple TBI studies, each with unique data-sharing restrictions. Access to the underlying data is available upon reasonable request to the principal investigators of each study.

## Acknowledgements

The authors gratefully acknowledge the patients and their families for contributing to the research.

