## Supplementary Material for "MRI improves global outcome prediction after moderate or severe traumatic brain injury"

**Table of Contents**

Supplementary Methods: pages 3-10

Supplementary Tables: pages 11-15

Supplementary Figures: pages 16-23

References: page 24-25

**Supplementary Methods**

External Validation Cohort

Between May 2024 and January 2026, we enrolled participants with moderate or severe TBI admitted to the Brigham and Women’s Hospital in Boston, MA, USA as part of an ongoing prospective study of TBI. Inclusion criteria were: age $\geq$ 18 years at time of admission, moderate or severe TBI, as defined by a Glasgow Coma Scale (GCS) score of < 13 documented in the hospital, when off sedation and in the absence of significant acute injury to other organ systems, abnormal brain CT scan, arrival to BWH within 2 weeks of injury, admitted at some point during hospital stay to an intensive care unit (ICU). Exclusion criteria were: moribund patient (expected not to survive 24 hours), pre-existing significant disability (modified Ranking Scale $\geq$ 3), inability to undergo MRI imaging. Multi-modal MRI was completed as soon as deemed safe by treating clinicians, and within 30 days of injury. Glasgow Outcome Scale Extended (GOSE) was collected a hospital discharge, 2-weeks, 6-months, and 1 year.

Primary Outcome

While the GOSE is traditionally measured without regard for the proximate cause of the resulting disability (whether due to the brain injury or trauma to other bodily systems),^1^ the GOSE-TBI is a new measure that scores disability specifically attributable to the brain injury, as opposed to other bodily trauma.^2^ This outcome measure was available only for a subset of the included studies (TRACK-TBI and RESPONSE-2). For consistency across datasets, we used the traditional GOSE. In the (N=4) instances where the GOSE was missing but GOSE-TBI was available, we used the GOSE-TBI.

### Image Processing

*Contusion tracing*

First, T1-weighted and FLAIR scans were screened for the presence/absence of contusions (W.H.C and S.B.S.; consensus review). Contusions were defined as hemorrhagic (T1 variable) and edematous (FLAIR hyperintense) lesions abutting a cortical surface and not conforming to a standard vascular distribution. The full extent of the contusion and surrounding edema was manually segmented (H.S. and J.L.S; ITKSnap, <https://github.com/pyushkevich/itksnap>) using the T1-weighted image to define the hemorrhagic core and the FLAIR scan to define the full lesion extent (Supplementary Figures 2,3). Lesions restricted to the white matter that did not reach the cortical surface were not traced. The resulting lesion masks were iteratively reviewed for accuracy (S.B.S.) and manually adjusted (H.S.). This procedure was repeated in the External Validation Cohort (M.G. and S.B.S). In all cases, segmentations were finalized while tracers were blinded to clinical outcomes.

Each participant’s T1-weighted sequence was registered to the standard Montreal Neurological Institute (MNI) T1 1mm template brain via rigid, affine, and deformable B-spline transformations (Advanced Normalization Tools [ANTs]^3–6^). Participants with severe anatomical distortions producing gross misalignment with the MNI T1 brain were excluded from analyses requiring contusion location (Supplementary Figure 1A).

*DTI cohort creation*

N=315 matched controls from the four studies were included for scaling participant FA values. To maximize power, within each study, each patient was compared against all controls scanned on the same scanner, even if there were slight differences in the acquisition protocol (e.g., voxel size, Repetition Time). As indicated in Supplementary Figure 1, for DTI-based analyses, participants were excluded for MRI acquired beyond 30 days post injury, severe anatomical distortions, inaccurate registrations to MNI space, or outlier amounts of motion (more extreme than any control).

DTI pre-processing included denoising (dwidenoise,^7^ MRITRIX) and eddy-current and motion correction (eddy^8^ from the FMRIB Software Library [FSL]). Fractional anisotropy (FA) maps were extracted with DTIFIT (FSL). Participant FA maps were registered to an MNI space FA template from 1,065 Human Connectome Project scans (HCP 1065),^9,10^ via rigid, affine and deformable B-spline symmetric normalization transformations (ANTs). Participants were excluded for excess motion (> 3.2 mm [most extreme control value]), or severe anatomical distortions preventing accurate registration into template space (Supplementary Figure 1B).

*Diffusion Tensor Imaging (DTI): White-matter skeleton*

Tract-Based Spatial Statistics (TBSS, FSL v6.0.7.11)^11^ was used to project voxel-wise FA values onto a cohort-wide consensus white-matter skeleton. We used the tbss_3_postreg and tbss_4_prestats commands from the Tract-Based Spatial Statistics (TBSS, FSL v6.0.7.11) software to generate a cohort-wide consensus white matter skeleton from individual FA maps registered into MNI space, which was then used to create a white-matter skeleton for each participant. Using the tbss_4_prestats command, the skeleton was thresholded at fractional anisotropy (FA) of 0.2.

*DTI region-of-interest (ROI) based approach*

To compare against skeleton lesion volume, we generated an alternate measure of white matter integrity, defined as the mean FA value within the white matter. We used the same FA maps that were registered to the Human Connectome Project standard FA map, as defined in the main text. We registered all ROIs from the Johns Hopkins University ICBM 1mm DTI white matter atlas (JHU ICBM;^12^ <https://identifiers.org/neurovault.image:1401>), excluding the fornix, to each participant’s native space using the previously generated transformations. After thresholding the FA maps at 0.2, we then extracted the mean FA of all nonzero voxels within a combined-mask of all white-matter ROIs. This represented the raw, unscaled FA values. Each raw value was then scaled by the control mean and standard deviation from the same scanner and study to obtain the “scaled” value.

*Diffusion Tensor Imaging (DTI): White-matter skeletons in External Validation Cohort*

DTI pre-processing in the External Validation Cohort followed the same denoising, eddy-current and motion correction as described for the Derivation Cohort, with the addition of *topup* applied to reverse-phase encoded field maps to unwarp magnetic field inhomogeneities. In N=7 scans where acquired multiband groups had unequal number of slices, blank slices were appended to the bottom to equalize slice numbers across groups as required by eddy software. Diffusion parameters were estimated with DTIFIT and registered to the same HCP-1065 FA template. Each new participant’s FA map was mapped directly onto the study-level skeleton from the Derivation Cohort; no new skeleton was generated. At the time of analysis, only 2 External Validation controls had been scanned at the Brigham and Women’s Hospital. To yield a group of controls adequate to estimate a mean and standard deviation at each point on the skeleton, we combined these two controls with N=13 controls scanned on the most similar (GE Signa 3T; Supplementary Table 1C) scanner from the TRACK TBI-study. Lesion maps along this skeleton were generated as in the Derivation cohort (2 SD below control mean). If a participant’s field of view excluded either the superior or inferior aspect of the Derivation Cohort skeleton, missing voxels were simply not included in the total lesion volume.

### Testing the prognostic value of Contusion/Axonal Injury location using the Damage Score

Among all participants with contusions, we performed voxel lesion symptom mapping (VLSM) within a leave-one-out cross-validation (LOOCV) framework using the Lesion to Symptom Mapping (LESYMAP, https://github.com/dorianps/LESYMAP) package in R (RStudio Version 2024.12.0+467). In each LOOCV iteration, we generated a t-statistic map of associations between lesion presence at each voxel (including voxels with ≥ 5 lesions) and 6-month GOSE. Cluster-level family-wise error correction was conducted using 1,000 permutations and a voxel-wise cluster-forming threshold of p < 0.001. Cluster significance (p < 0.05) was determined by comparing observed cluster sizes to the maximum-cluster-size null distribution.

A Damage Score (Supplementary Figure 3) was then computed for each held-out lesion, representing the total degree of overlap between the held-out lesion and the significant clusters from the VLSM map. Using a previously published approach, ^13–15^ we defined the Damage Score as the sum of the map’s statistical values falling inside the held-out lesion. This procedure weights the overall lesion volume such that a lesion overlapping areas of stronger association will have a higher score than an equivalently sized lesion outside these areas. Finally, we tested the hypothesis that the iteratively computed Damage Score has a different discriminative performance (AUC) than contusion volume using Delong’s test.

In the same manner as described for contusions, we performed VLSM within a LOOCV framework to generate maps of statistical association between lesion presence at every voxel along the consensus white-matter skeleton and GOSE outcomes (Supplementary Figure 4). The held-out participants’ axonal injury lesion masks were used as above to compute a Damage Score. The AUCs of the iteratively generated Damage Score and axonal injury lesion volume were compared using Delong’s test.

### Connectome Analyses

Because the structural and functional connectivity of lesion locations has been shown to be important for explaining symptoms,^16^ we repeated the above-described VLSM and damage score procedure using brain-wide maps of voxels structurally and functionally connected to the contusion location in large normative connectomes.

*Contusion structural disconnection mapping*

To explore this, we used the Brain Connectivity Behavior (BCBToolkit, http://toolkit.bcblab.com/)^17^ Disconnectome 7T, a 178-subject dataset with 7T, 1 mm imaging data from the Human Connectome Project. Each participant’s contusion mask was overlaid onto the tractography map. The percentage of streamlines passing through each lesion was retained to create a white-matter disconnection map. Disconnection maps were then thresholded at 0.5 and binarized, leaving only voxels where more than 50% of streamlines ultimately crossed participant’s contusion mask. We used the LESYMAP software to compute the voxelwise associations between disconnection maps and 6-month GOSE outcomes within a leave one-subject out framework, with an FDR multiple comparisons correction threshold applied to each map at 0.05. The resulting thresholded t-statistic map of associations with GOSE was then multiplied by the held-out contusion mask (not used to generate the association map). The t values inside the contusion mask were then summed to compute the Damage Score.

*Contusion functional network mapping*

A gender-balanced Brain Genomics Superstruct Project (GSP)^18,19^ connectome with 1000 participants was used to compute the seed-to-voxel functional connectivity between participants’ contusion mask and every other brain voxel in MNI T1 2 mm space, using standard lesion network mapping methodologies.^16^ The resulting map describes the strength of correlation in the brain-oxygen level dependent (BOLD) signal between the location of the lesion and every other brain voxel. The t-statistic map was created from lesion network maps using randomise (FSL)^20^, a similar voxel-wise analysis-based software. As with the leave-one-out approach described in the main text, the “Damage Score” was created from the product of the unthresholded t-statistic map and the held-out contusion label. To accommodate the standard voxel size, in the normative functional connectivity data, contusion labels were down-sampled to 2mm from 1mm space.

*Axonal injury structural disconnection mapping*

Using a similar method as described for cohort 1, we calculated the map of white matter tracts in a normative connectome intersecting each participant’s axonal injury lesion mask. As above, the disconnection maps were thresholded at 0.5 and binarized. Within a leave-one out framework, LESYMAP was again used to compute the voxel-wise association between disconnection maps and 6-month GOSE outcomes (cluster-level P_FWE_ < 0.05). The Damage Score was computed as the sum of association-map t values inside the held-out participant’s axonal injury lesion mask.

**Supplementary Tables**

| **Supplementary Table 1A:** BIOAX DTI Acquisition Parameters | | | | | | | |
| --- | --- | --- | --- | --- | --- | --- | --- |
| **BIOAX BIDS** | | | | | | | |
| Scanner | Siemens Skyra 3T | Philips Achieva  1.5T | Philips Achieva 3T | Siemens Aera 1.5T | GE Optima 1.5T | Siemens TrioTim 3T | Siemens Verio 3T |
| Number of Subjects | Ctrl: 14  Pt:1 | 14  10 | 14  4 | 13  5 | 15  16 | 14  7 | 33  4 |
| Diffusion directions | 64 | 32 | 64 | 54 | 60 | 64 | 64 |
| Gradient Strength | 1000 | 1000 | 1000 | 1000 | 1000 | 1000 | 1000 |
| B_0_ volumes | 3 | 1 | 4 | 1 | 1 | 1 | 4 |
| Voxel size (mm) | 2x2x2 | 2x2x2 | 1.6x1.6x2 | 2x2x2 | 1x1x2 | 2x2x2 | 2x2x2 |
| Repetition Time (s) | 5.1 | 9.5 | 9.5 | 12.3 | 15.5 | 9.5 | 9.5 |
| Echo Time (s) | 0.1 | 0.103 | 0.103 | 0.081 | 0.104 | 0.103 | 0.103 |
| Multiband | 2 | 1 | 1 | 1 | 1 | 1 | 1 |

| **Supplementary Table 1B:** RESPONSE Pilot and RESPONSE 2 DTI Acquisition Parameters | | | | |
| --- | --- | --- | --- | --- |
| **RESPONSE Pilot** | | | **RESPONSE 2** | |
| Scanner | Siemens Skyra 3T | Siemens Skyra 3T | Scanner | Siemens Skyra 3T |
| Number of Subjects | Ctrl: 12  Pt: 8 | 4  7 | Number of Subjects | 28  29 |
| Diffusion directions | 60 | 60 | Diffusion directions | 60 |
| Gradient Strength | 2000 | 2000 | Gradient Strength | 2000 |
| B_0_ volumes | 10 | 10 | B_0_ volumes | 10 |
| Voxel size (mm) | 2x2x2 | 2x2x2 | Voxel size (mm) | 2x2x2 |
| Repetition Time (s) | 6.8 | 13.7 | Repetition Time (s) | 6.7 |
| Echo Time (s) | 0.097 | 0.098 | Echo Time (s) | 0.1 |

| **Supplementary Table 1C:** TRACK-TBI Philips Achieva and GE Signa DTI Acquisition Parameters | | | | |
| --- | --- | --- | --- | --- |
| **TRACK-TBI** | | | | |
| **Philips Achieva 3T** | | | **GE Signa 3T** | |
| Protocol | Protocol 1 | Protocol 2 | Protocol | Protocol 1 |
| Number of Subjects | Ctrl: 17  Pt: 16 | 8  1 | Number of Subjects | 13  1 |
| Diffusion directions | 64 | 64 | Diffusion directions | 64 |
| Gradient Strength | 1300 | 1300 | Gradient Strength | 1300 |
| B_0_ volumes | 1 | 1 | B_0_ volumes | 8 |
| Voxel size (mm) | 2.73x2.73x2.70 | 2.73x2.73x2.70 | Voxel size (mm) | 1.33x1.33x2.70 |
| Repetition Time (s) | 9.00 | 9.00 | Repetition Time (s) | 15.50 |
| Echo Time (s) | 0.071 | 0.094 | Echo Time (s) | 0.087 |

| **Supplementary Table 1D:** TRACK-TBI GE Discovery DTI Acquisition Parameters | | | |
| --- | --- | --- | --- |
| **GE Discovery 3T** | | | |
| Protocol | Protocol 1 | Protocol 2 | Protocol 3 |
| Number of Subjects | Ctrl: 27  Pt: 6 | 9  1 | 0  5 |
| Diffusion directions | 64 | 64 | 64 |
| Gradient Strength | 1300 | 1000 | 1300 |
| B_0_ volumes | 8 | 1 | 8 |
| Voxel size (mm) | 1.37x1.37x2.70 | 0.90x0.90x2.40 | 1.37x1.37x2.70 |
| Repetition Time (s) | 9.05 | 3.00 | 9.00 |
| Echo Time (s) | 0.081 | 0.094 | 0.082 |

| **Supplementary Table 1E:** TRACK-TBI Siemens Triotim DTI Acquisition Parameters | | | | |
| --- | --- | --- | --- | --- |
| **Siemens Triotim 3T** | | | | |
| Sequence | Protocol 1 | Protocol 2 | Protocol 3 | Protocol 4 |
| Number of Subjects | Ctrl: 25  Pt: 3 | 8  10 | 10  4 | 0  9 |
| Diffusion directions | 64 | 64 | 63 | 64 |
| Gradient Strength | 1300 | 1300 | 1000 | 1000 |
| B_0_ volumes | 1 | 1 | 6 | 5 |
| Voxel size (mm) | 2.73x2.73x2.70 | 2.73x2.73x2.63 | 2.40x2.40x2.40 | 2.40x2.40x.2.40 |
| Repetition Time (s) | 9.00 | 9.00 | 2.90 | 3.70 |
| Echo Time (s) | 0.094 | 0.094 | 0.094 | 0.092 |

| **Supplementary Table 1F:** TRACK-TBI Siemens Skyra DTI Acquisition Parameters | | | | | | | |
| --- | --- | --- | --- | --- | --- | --- | --- |
| **Siemens Skyra 3T** | | | | | | | |
| Protocol | Protocol 1 | Protocol 2 | Protocol 3 | Protocol 4 | Protocol 5 | Protocol 6 | Protocol 7 |
| Number of Subjects | Ctrl: 21  Pt: 1 | 1  11 | 1  7 | 9  5 | 1  0 | 2  0 | 2  3 |
| Diffusion directions | 64 | 64 | 64 | 64 | 64 | 64 | 64 |
| Gradient Strength | 1000 | 1000 | 1000 | 1300 | 1300 | 1000 | 1000 |
| B_0_ volumes | 6 | 6 | 5 | 8 | 1 | 6 | 6 |
| Voxel size (mm) | 2.40x1.40x1.40 | 2.40x2.40x2.40 | 2.40x2.40x2.40 | 2.73x2.73x2.70 | 2.73x2.73x2.70 | 2.50x2.40x2.40 | 2.40x2.40x2.40 |
| Repetition Time (s) | 2.90 | 2.90 | 2.90 | 9.00 | 9.00 | 3.70 | 2.90 |
| Echo Time (s) | 0.12 | 0.094 | 0.094 | 0.092 | 0.094 | 0.092 | 0.092 |

| **Supplementary Table 1G:** External Validation Cohort DTI Acquisition Parameters | | | | |
| --- | --- | --- | --- | --- |
| **Brigham and Women’s** | | | | |
| Scanner | GE Signa Architect 3T | GE Signa Architect 3T | GE Signa Architect 3T | GE Signa Architect 3T |
| Number of Subjects | Ctrl: 2  Pt: 21 | Ctrl: 0  Pt: 5 | Ctrl: 0  Pt: 1 | Ctrl: 0  Pt: 1 |
| Diffusion directions | 64 | 64 | 64 | 50 |
| Gradient Strength | Multi-shell: 1000, 2000 | Multi-shell: 1000, 2000 | Multi-shell: 1000, 2000 | Multi-shell: 1000, 2000 |
| B_0_ volumes | 11 | 11 | 11 | 7 |
| Voxel size (mm) | 2x2x2 | 1.981x1.981x2 | 2.264x2.264x2 | 2x2x2 |
| Repetition Time (s) | 5.269 - 8.118 | 6-8.967 | 6.2 | 5.8 |
| Echo Time (s) | 0.0897-0.1107 | 0.0898-0.1153 | 0.0894 | 0.1015 |

| **Supplementary Table 2:** Demographic Data Across Studies (N=201) | | | | | |
| --- | --- | --- | --- | --- | --- |
| Characteristics | BIOAX  (N=65) | RESPONSE 2  (N=31) | RESPONSE Pilot  (N=18) | TRACK  (N=87) | *P* Value |
| Mean Age (SD) | 46 (18) | 44 (21) | 29 (9) | 38 (18) | 0.0022 |
| Male Sex (%) | 55/65 (85%) | 22/31 (71%) | 13/18 (72%) | 56/87 (64%) | 0.047 |
| Race (%)  White  Black  Other | 59/64 (92%)  3/64 (5%)  2/64 (3%) | 24/28 (86%)  3/28 (11%)  1/28 (4%) | 15/18 (83%)  0/18 (0%)  3/18 (17%) | 73/85 (86%)  9/85 (11%)  3/85 (4%) | 0.19 |
| Median Admission GCS (IQR) | 7 (4) | 7 (5) | 6 (2) | 9 (9) | 0.08 |
| Pupils (%)  Both React  One Reacts  Neither Reacts | 52/61 (85%)  8/61 (13%)  1/61 (2%) | 23/29 (79%)  3/29 (10%)  3/29 (10%) | 10/17 (59%)  1/17 (6%)  6/17 (35%) | 67/77 (87%)  1/77 (1%)  9/77 (12%) | 0.001 |
| CT Marshall Score (%)  I  II  III  IV  V  VI | 5/62 (8%)  27/62 (44%)  5/62 (8%)  2/62 (3%)  4/62 (7%)  19/62 (31%) | 0/31 (0%)  13/31 (42%)  7/31 (23%)  0/31 (0%)  5/31 (16%)  6/31 (19%) | 0/18 (0%)  7/18 (39%)  8/18 (44%)  0/18 (0%)  3/18 (17%)  0/18 (0%) | 32/75 (43%)  37/75 (49%)  2/75 (35)  3/75 (4%)  0/75 (0%)  1/75 (1%) | <0.001 |
| Hemorrhagic Contusion (%) | 47/65 (72%) | 26/31 (84%) | 17/18 (94%) | 49/87 (56%) | 0.001 |
| Median Hemorrhagic Contusion Volume (cc, IQR) | 22 (67) | 44 (85) | 38 (56) | 2 (18) | <0.001 |
| Median Axonal Injury Grade (IQR) | 2 (2) | 2 (1) | 2 (2) | 2 (2) | 0.11 |
| Mean Scaled White Matter Mean FA (SD) | -1.60 (1.36) | -2.50 (1.68) | -2.87 (1.32) | -0.88 (1.12) | <0.001 |
| Median White Matter Lesion Volume (cc, IQR) | 6.67 (9.58) | 17.06 (15.23) | 16.31 (13.56) | 3.77 (5.24) | <0.001 |
| Median Days to MRI (IQR) | 21 (10) | 10 (8) | 10 (5) | 14 (0) | <0.001 |
| Mean (mm) Motion During DTI (SD) | 0.82 (0.82) | 0.94 (0.49) | 1.90 (1.80) | 0.80 (0.61) | <0.001 |
| 6-month GOSE $\leq$ 4 (%) | 31/65 (48%) | 20/31 (65%) | 9/18 (50%) | 15/87 (17%) | <0.001 |

| **Supplementary Table 3:** Patient Characteristics | | | |
| --- | --- | --- | --- |
|  | Derivation Cohort (N=201) | External Validation Cohort  (N = 27) | **p-value** |
| Mean Age (SD) | 41 (18) | 59 (22) | < 0.0001 |
| Male Sex (%) | 146/201 (73%) | 22/27 (81%) | 0.33 |
| Race (%)  White  Black  Other | 171/195 (88%)  15/195 (8%)  9/195 (5%) | 19/27 (70%)  2/27 (7%)  6/27 (22%) | 0.003 |
| Mean Admission GCS (SD) | 7 (4) | 6 (3) | 0.18 |
| Pupils (%)  Both React  One Reacts  Neither Reacts | 152/184 (83%)  13/184 (7%)  19/184 (10%) | 19/26 (73%)  2/26 (8%)  5/26 (19%) | 0.40 |
| CT Marshall Score (%)  I  II  III  IV  V  VI | 37/186 (20%)  84/186 (45%)  22/186 (12%)  5/186 (3%)  12/186 (6%)  26/186 (14%) | 0/27 (0%)  14/27 (52%)  3/27 (11%)  1/27 (4%)  3/27 (11%)  6/27 (22%) | 0.19 |
| Hemorrhagic Contusion (%) | 138/200 (69%) | 25/27 (93%) | 0.01 |
| Median Hemorrhagic Contusion Volume (cc, IQR) | 11 (59) | 28 (110) | 0.05 |
| Median White Matter Lesion Volume (cc, IQR) | 6.6 (12.8) | 13.0 (8.3) | <0.001 |
| Median Days to MRI (IQR) | 14 (2) | 7 (8) | <0.001 |
| Mean (mm) Motion During DTI (SD) | 0.93 (0.90) | 1.11 (0.69) | 0.24 |
| Median IMPACT_Core+CT+Lab_ Probability of Unfavorable Outcome (IQR) | 0.37 (0.38) | 0.58 (0.33) | 0.002 |
| 6-month GOSE $\leq$ 4 (%) | 75/201 (37%) | 14/27 (52%) | 0.21 |

| **Supplementary Table 4: Multivariate Model for Unfavorable 6-month Outcome** | | | |
| --- | --- | --- | --- |
|  | Coefficient | Adjusted Odds Ratio | P value |
| Intercept | -6.1 | -- | < 0.001 |
| Age (per 10 years) | 0.38 | 1.5 [1.2, 1.8] | 0.001 |
| Log2 Contusion Volume | 0.28 | 1.3 [1.1, 1.6] | 0.002 |
| Log2 Axonal Injury Volume | 0.85 | 2.3 [1.5, 3.6] | < 0.001 |

**Supplementary Figures**

**Supplementary Figure 1: Analytic Cohort CONSORT Diagrams**

Consort diagrams demonstrating the specific cohort analyzed in each of the main text figures. These cohorts varied slightly due to the different imaging requirements. The External Validation Cohort is shown in (D). MRI (Magnetic Resonance Imaging); DTI Diffusion Tensor Imaging; WLST withdrawal of life sustaining therapy

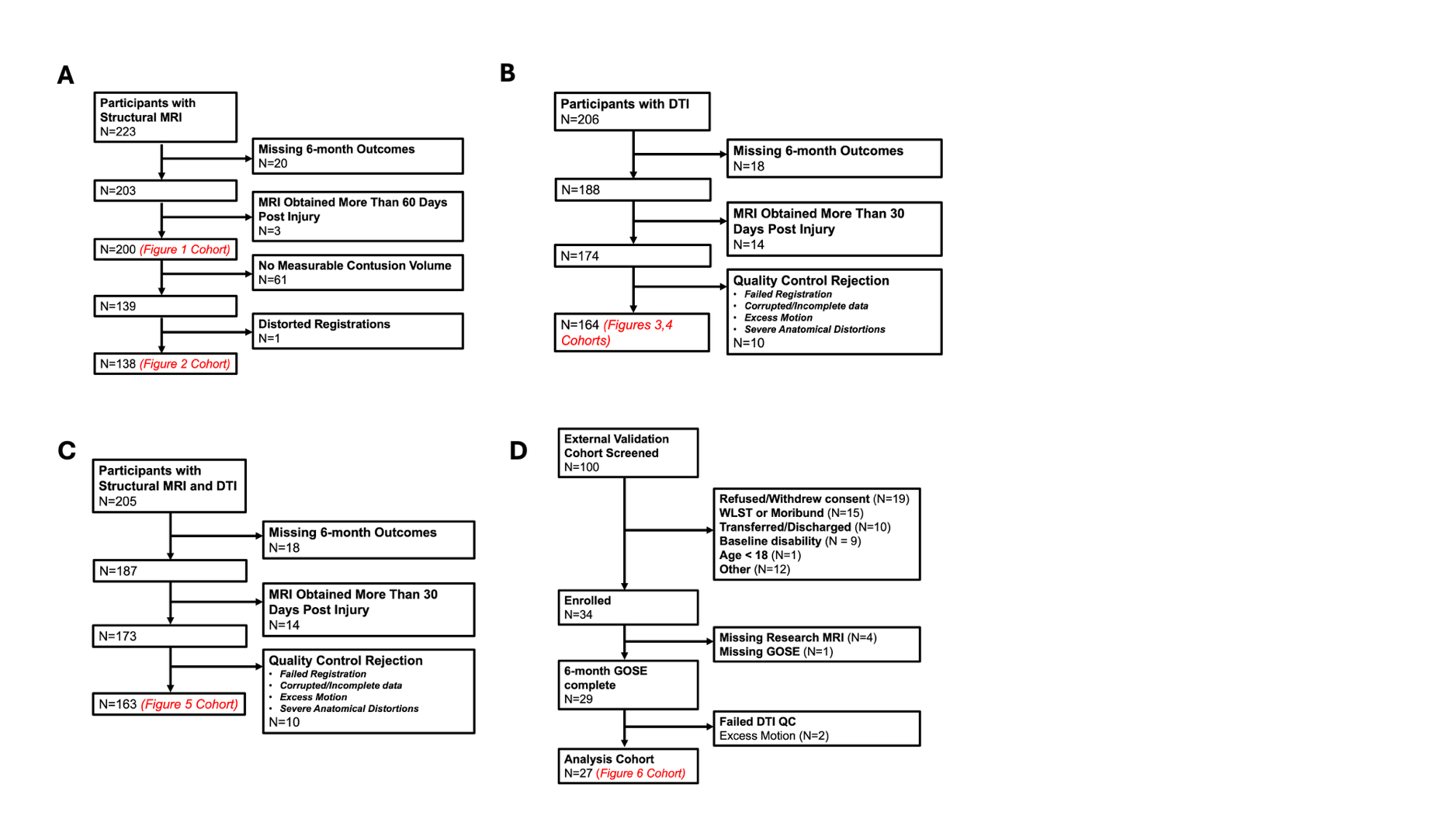

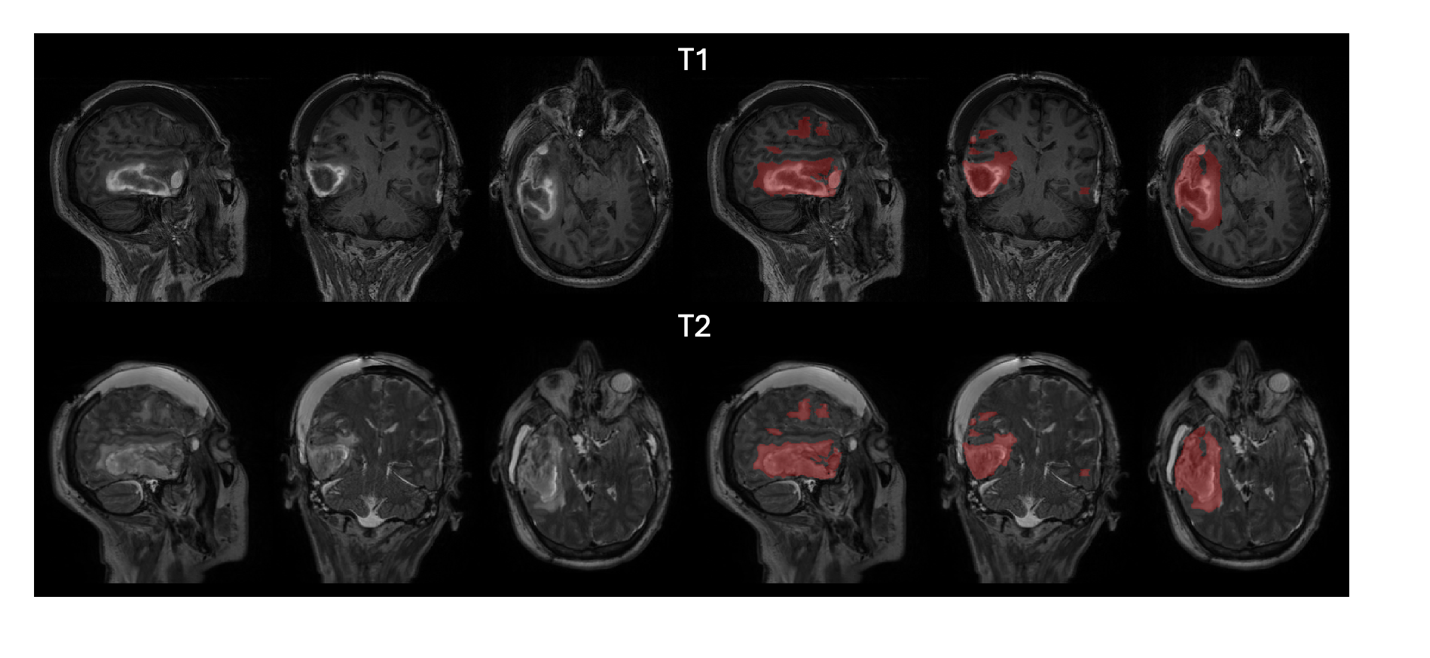

**Supplementary Figure 2 An example of a contusion segmentation**

The area of hemorrhage was identified on the T1-weighted sequence (T1-variable) and surrounding edema on the T2-weighted sequence (T2-bright).

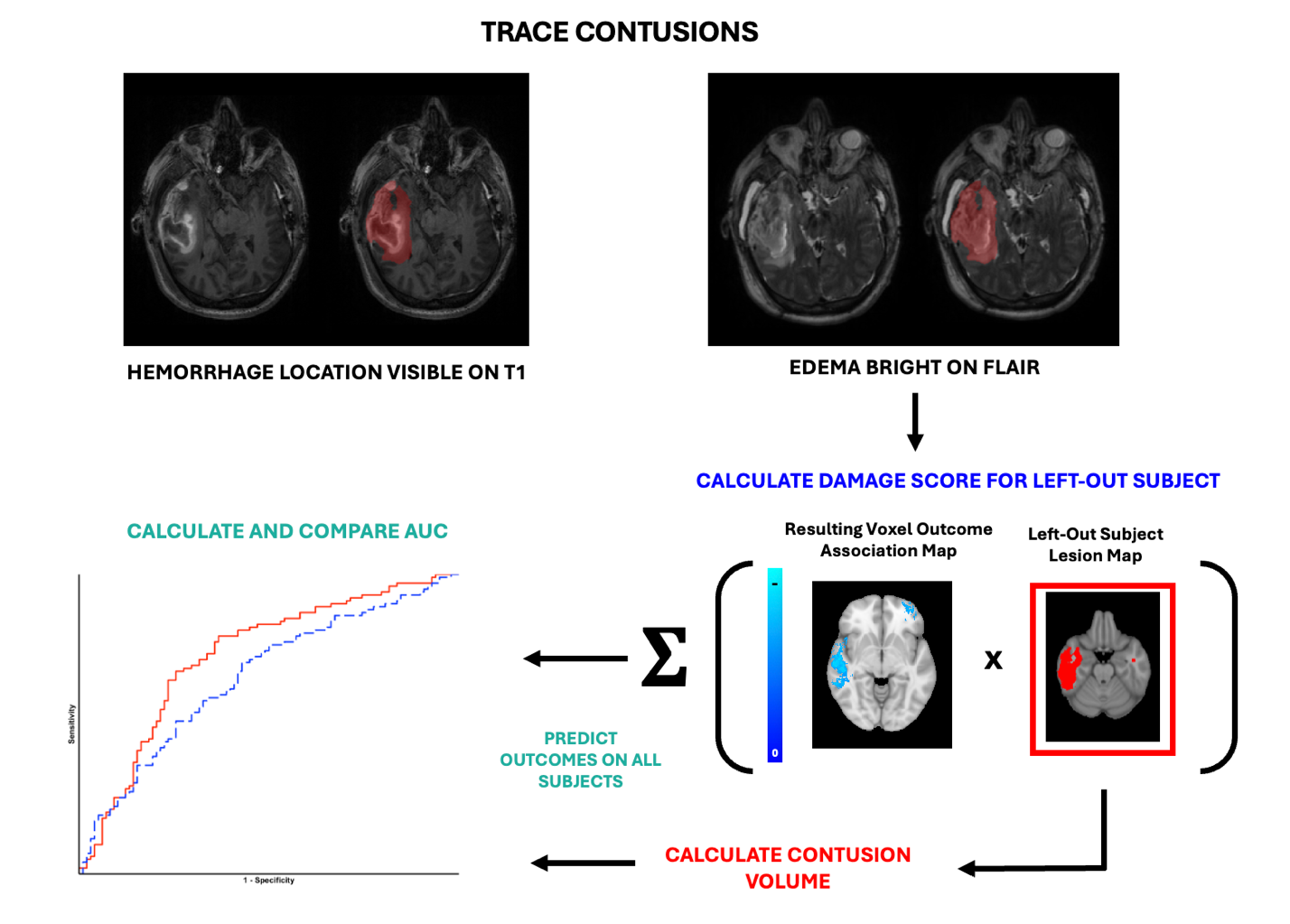

**Supplementary Figure 3: Methods schematic for testing the discriminative accuracy of contusion volume and location**

Lesion masks were traced for each participant in the cohort based on T1 and T2-weighted imaging. Contusion lesion masks were then registered into MNI 152 T1 1mm space. A leave one out cross validation (LOOCV) was performed to produce a location-weighted contusion volume metric (Damage Score) by multiplying the held-back contusion mask by a map of statistical associations with outcome generated in the remaining data. We then compared the Damage Score (weighted by lesion location) and unweighted (total lesion volume alone) measures in discriminative accuracy using area under the receiver operating characteristic curve.

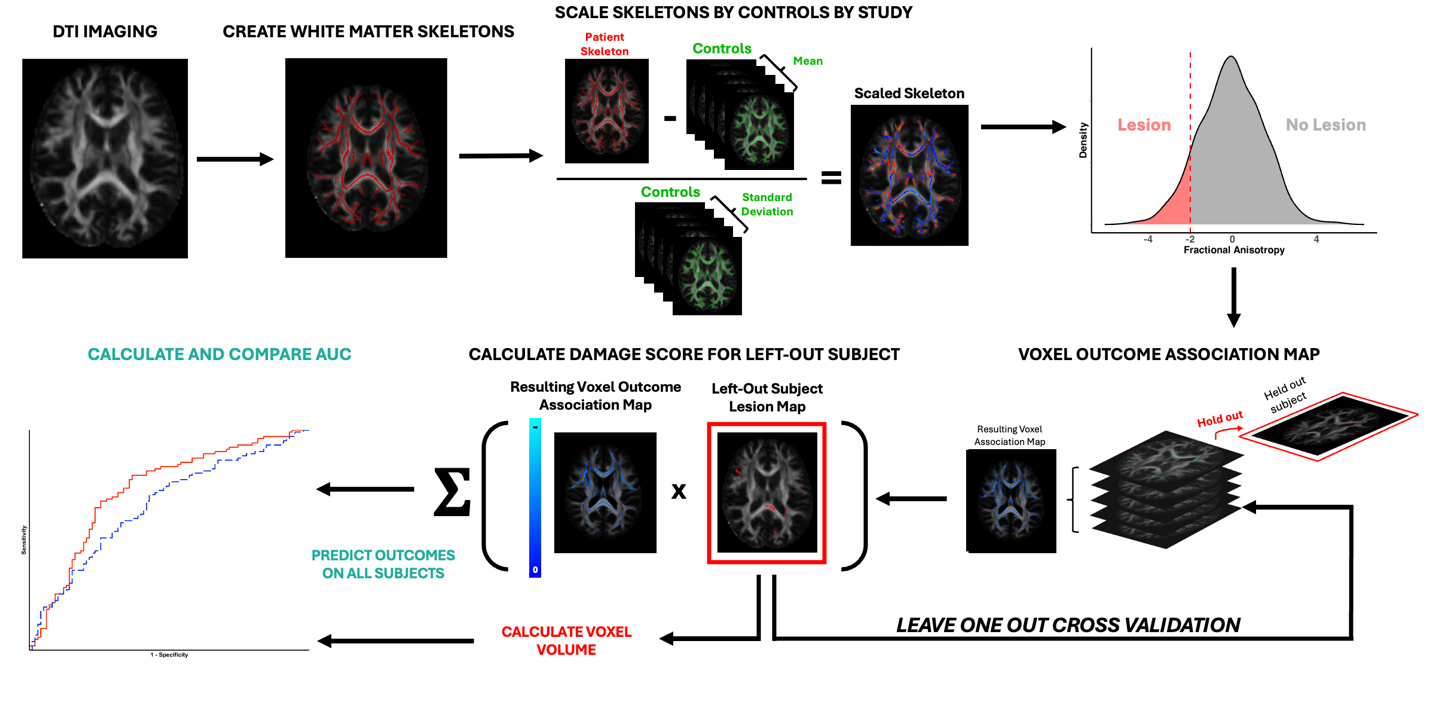

**Supplementary Figure 4: Methods Schematic for testing the discriminative accuracy of axonal injury volume and location**

DTI was preprocessed into fractional anisotropy maps, which were compared to control maps using an approach similar to one that has been previously described (Jolly et. al, 2020). We defined lesions based on fractional anisotropy along a white matter skeleton at least 2 standard deviations below the control mean. We performed a leave one out cross validation (LOOCV), multiplying the held-out mask by the map of statistical associations with outcome generated in the remaining data, to generate the Damage Score (location dependent). We then compared the discriminative accuracy of this metric to the total volume of lesioned voxels along the white matter skeleton.

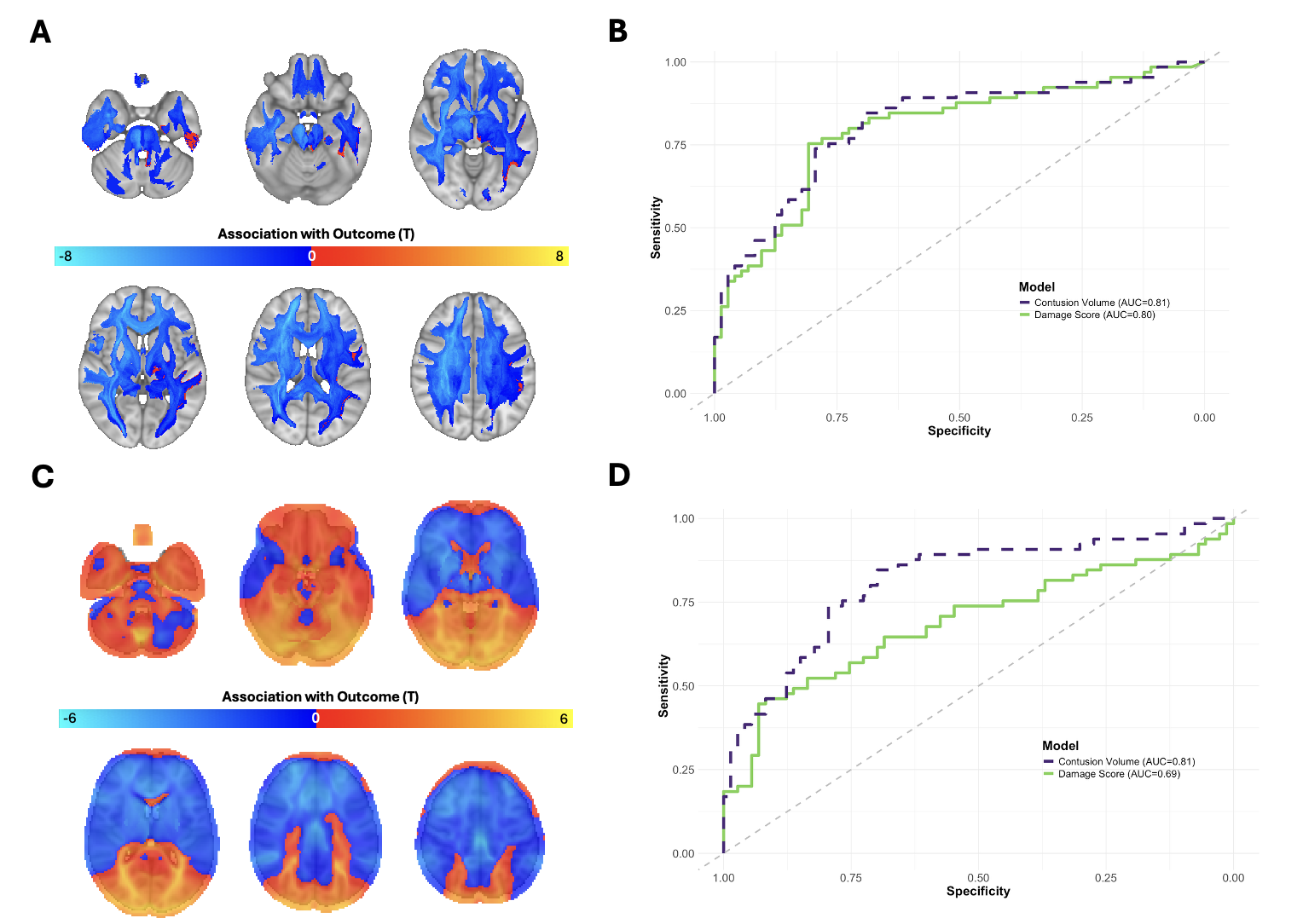

**Supplementary Figure 5: The normative, brain-wide structural and functional connectivity of contusions do not improve outcome discrimination**

(A) T-statistic maps generated using structural disconnection maps showing association with 6-month GOSE outcomes. Areas that are negative are those that when disrupted result in a lower 6-month GOSE scoring (B) ROC curves comparing structural connectivity derived Damage Scores (green) and contusion volume (purple). (C) T-statistic maps generated using contusion functional connectivity maps. Areas that are negative are those where higher functional connectivity values are associated with lower 6-month GOSE scores. (D) ROC curves comparing functional connectivity Damage Score (green) with contusion volume (purple).

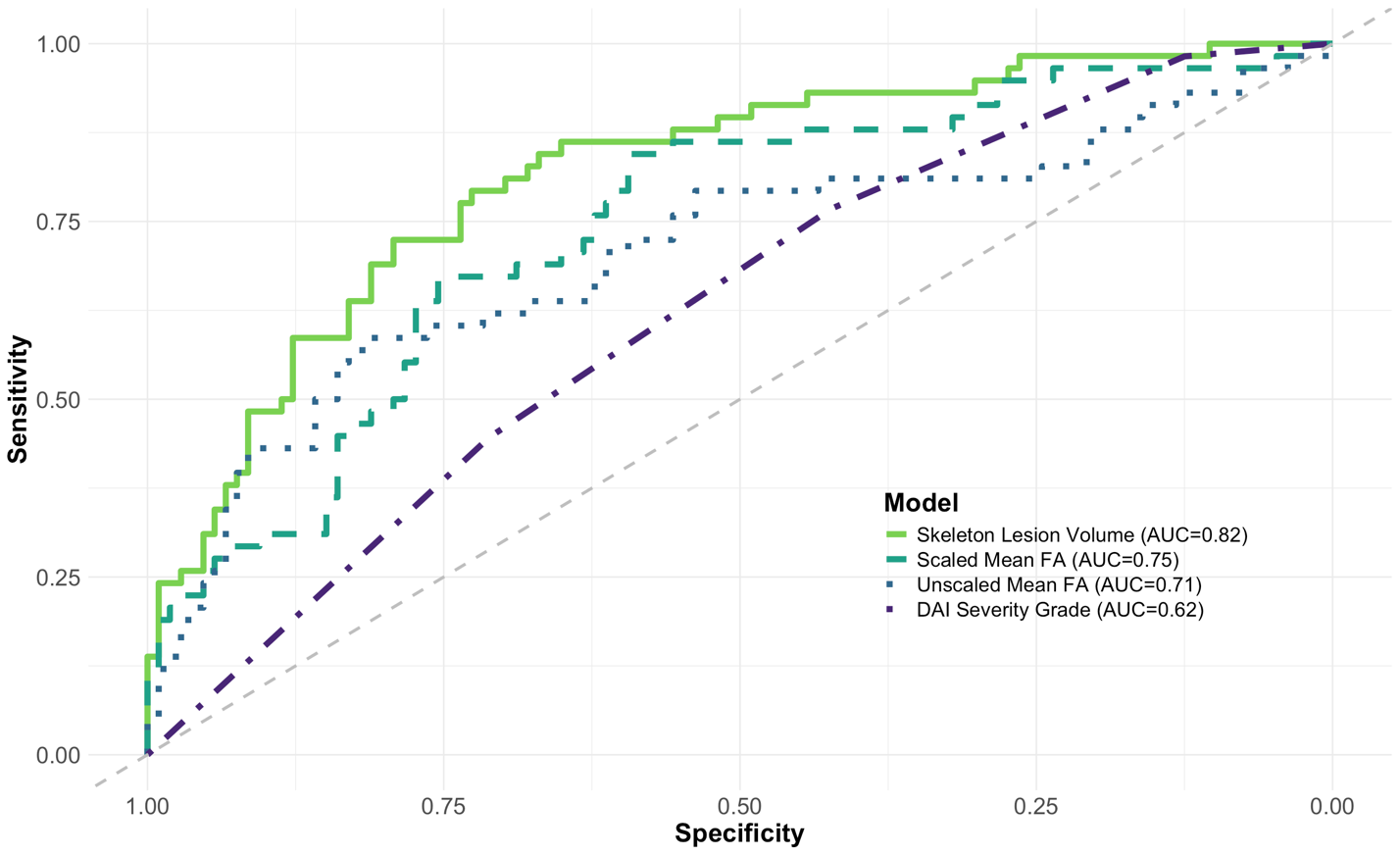

**Supplementary Figure 6: Comparison of different approaches to measuring axonal injury**

Axonal injury volume measured as volume of voxels with FA more than 2 SD below control mean on a standard white matter skeleton (solid), mean FA within a standard white matter atlas, scaled by scanner controls (dots), mean FA within a standard white matter atlas with no control scaling (dashed), and visual Adams/Gentry DAI grade based on standard structural MRI sequences (dot/dash).

**Supplementary Figure 7: Brain-wide normative structural connectivity of axonal injury locations does not improve outcome discrimination.** A) Voxel-wise association T-statistic map. Only voxels that survived permutation-based cluster multiple comparisons correction (raw p threshold of 0.001; P_FWE_ < 0.05 voxels are shown. B) ROC curves comparing the discriminative ability of structural-connectivity based Damage Score (green) to axonal injury severity (purple).

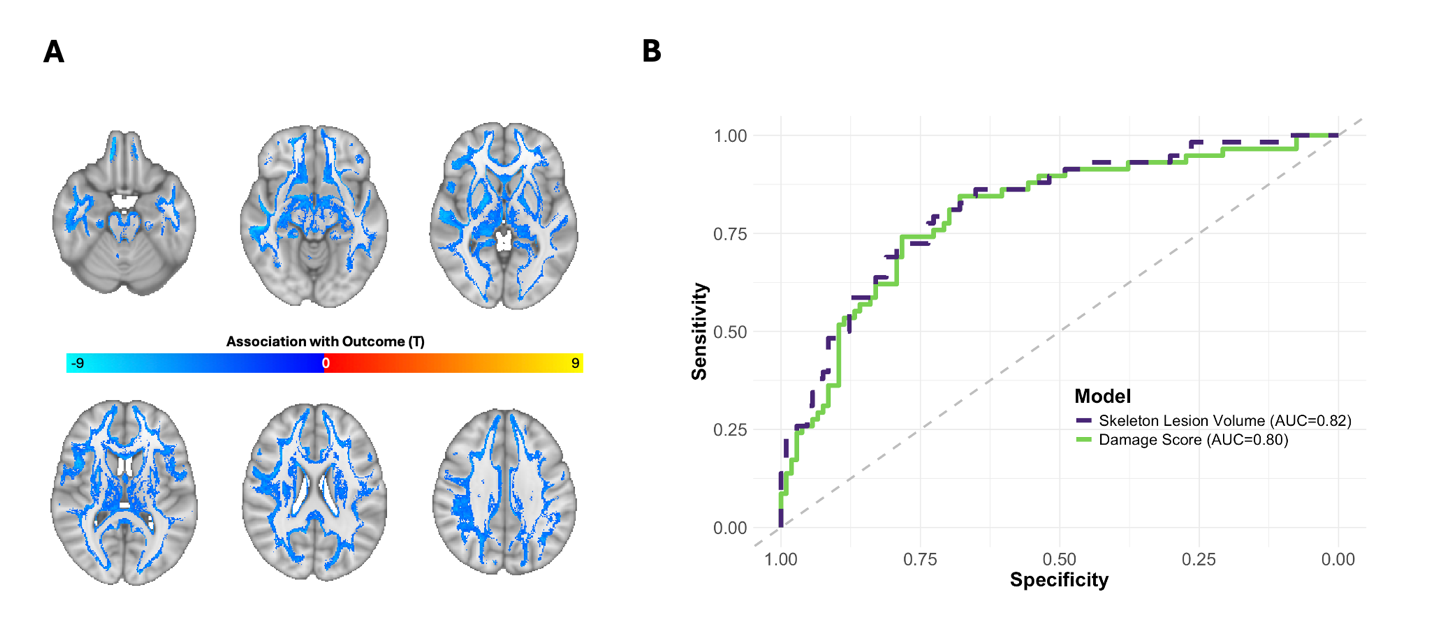

**Supplementary Figure 8: MRI-measured lesion volumes improve the IMPACT model** Validation Cohort ROC curves for IMPACT models fit (inside the Derivation Cohort) with all existing variables (IMPACT_Core+Extended+Labs_) or with CT-based variables replaced with MRI-measured lesion volumes (IMPACT_Core+MRI+Labs_).

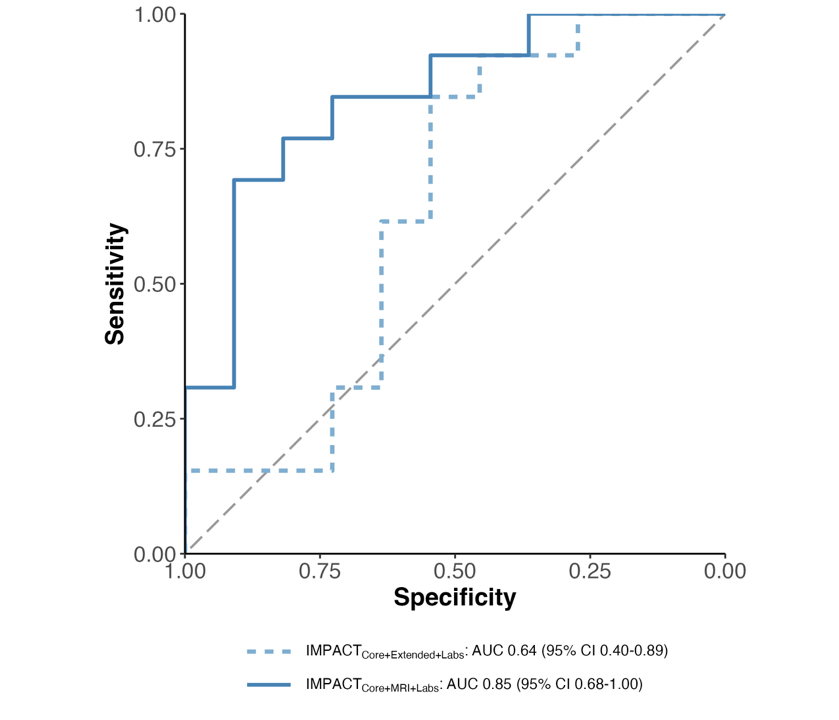
